# Airway Spatial Transcriptomics in Smoking

**DOI:** 10.1101/2025.04.01.25325047

**Authors:** Jarrett D. Morrow, Zaid W. El-Husseini, Jeong H. Yun, Craig P. Hersh

## Abstract

**Background:** Cigarette smoking has a significant impact on global health. Although cessation has positive health benefits, some molecular changes to intercellular communications may persist in the lung. In this study we created a framework to generate hypotheses by predicting altered cell-cell communication in smoker lungs using single-cell and spatial transcriptomic data.

**Methods:** We integrated publicly available lung single-cell transcriptomic data with spatial transcriptomic data from never-smoker and current-smoker lung tissue samples to create spatial transcriptomic data at virtual single-cell resolution by mapping individual cells from our lung scRNA-seq atlas to spots in the spatial transcriptomic data. Cell-cell communications altered in smoking were identified using the virtual single-cell transcriptomic data.

**Results:** We identified pathways altered in the three current-smoker samples compared with the three never-smoker samples, including the up-regulated collagen pathway. We observed increased collagen pathway activity involving the ligands COL1A1 and COL1A2 in adventitial fibroblasts and decreased activity involving COL1A2 and COL6A3 in pericytes and myofibroblasts, respectively. We also identified other pathways with structural (e.g. Fibronectin-1), immune-related (e.g. MHC-II), growth factor (e.g. Pleiotrophin) and immunophilin (e.g. Cyclophilin A) roles.

**Conclusions:** In this study we inferred spatially proximal cell-cell communication between interacting cell types from spatial transcriptomics at virtual single-cell resolution to identify lung intercellular signaling altered in smoking. Our findings further implicate several pathways previously identified, and provide additional molecular context to inform future functional experiments and therapeutic avenues to mitigate pathogenic effects of smoking.

## Introduction

Cigarette smoking has a significant impact on global health, with 480,000 smoking-related deaths in the United States each year [1]. Smoking is major driver of morbidity and mortality and a significant risk factor for several lung diseases, including chronic obstructive pulmonary disease (COPD) and idiopathic pulmonary fibrosis (IPF). Approximately 28.8 million adults in the United States smoked cigarettes in 2022 [2], with 8.8% successfully quitting. Although cessation has positive health benefits [3,4], some molecular changes due to smoking may persist in the lung [5–12].

Cell-cell Interactions are physical (e.g. cell adhesion) or biochemical (e.g. ligand from one cell binding to receptor on another) interactions between cells, with key roles in the function of tissues [13–15]. Although not studies of smoking, Blackburn et al. sought to identify lung cell-cell interactions altered in COPD using scRNA-seq data [16] and Browaeys et al. compared healthy lungs to IPF lungs using scRNA-seq to detect disease-relevant cell-cell communications [17].

Spatial transcriptomic technologies provide the ability to examine cell-cell communication with increased rigor, leveraging information regarding cell locations and proximity [18]. Spatial transcriptomic profiling at single-cell resolution is possible (e.g. 10x Genomics Visium HD or Xenium). However, in this study we sought to leverage publicly available data from lower-cost technologies across several lung tissue studies. We developed a framework to integrate single-cell and spatial transcriptomic data and predict lung cell-cell communication. A better understanding of intercellular signaling altered in smoker lungs may inform therapeutic development efforts to mitigate the pathogenic effects of smoking [13,14].

## Methods

Lung spatial transcriptomic data at virtual single-cell resolution were created using the cellular (Cyto) Spatial Positioning Analysis via Constrained Expression alignment (CytoSPACE) method to efficiently map individual cells from a lung scRNA-seq atlas to spots in lung spatial transcriptomic data from never-smoker (NS) and current-smoker (CS) lung tissue samples (Figure 1A) [19]. Cell-cell communications altered by smoking were identified using CellChat with these spatial transcriptomic data [20].

**Figure 1.**
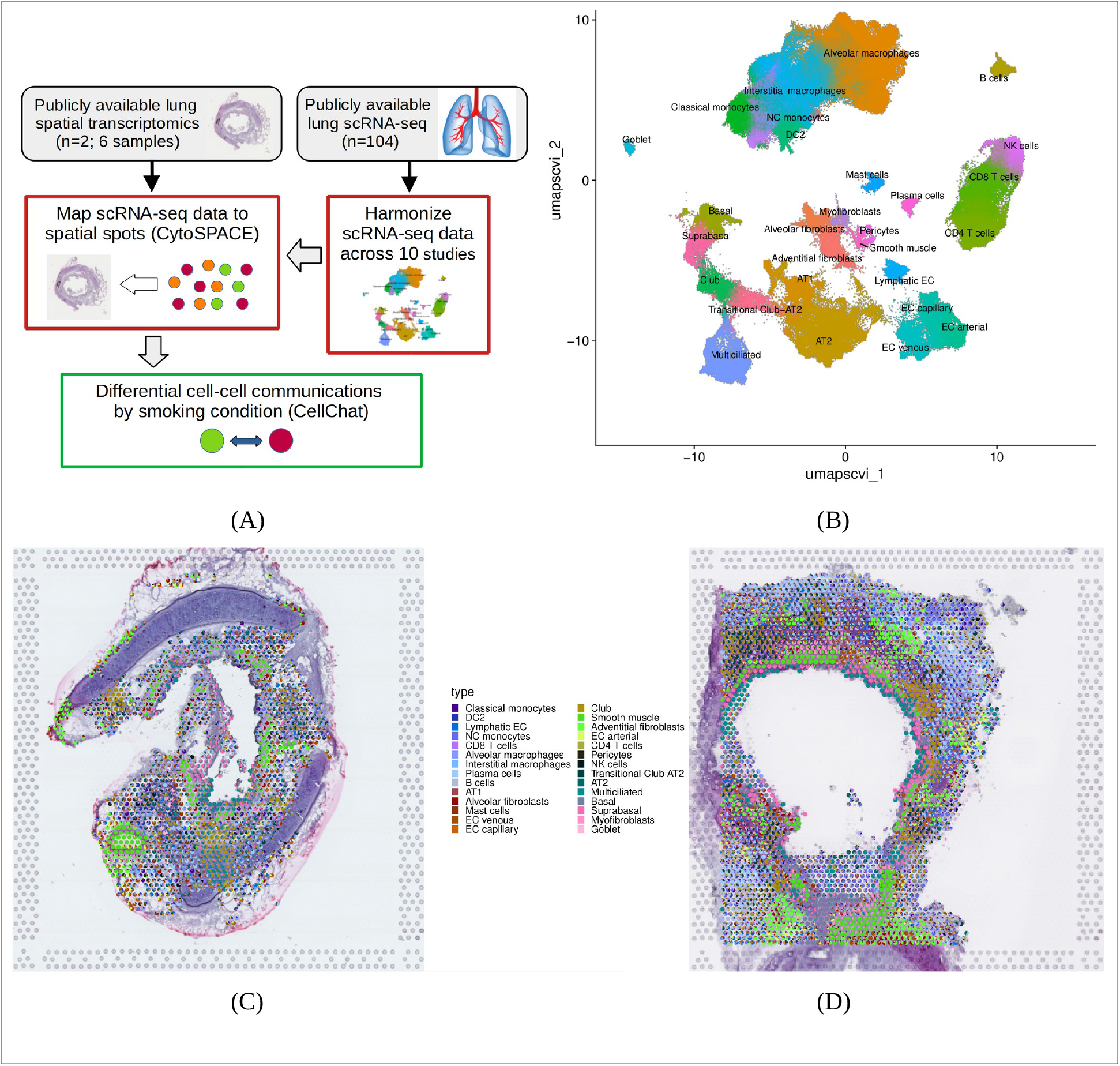
(A) The study involved use of bioinformatic methods with publicly available lung single-cell and spatial transcriptomic data. The single-cell RNA-seq data were harmonized to create a single-cell reference dataset for use with CytoSPACE. (B) Each cell-type identified across the ten studies shown in the UMAP plot. CytoSPACE maps individual cells from the scRNA-seq dataset onto each spot within the spatial transcriptomic data. The cell-type composition for a never-smoker sample (C; WSA_LngSP8759312) and a current-smoker sample (D; WSA_LngSP9258464).

### Lung single-cell RNA-seq data

The lung scRNA-seq reference atlas for use with CytoSPACE was created by harmonizing data from ten prior lung tissue studies (n=104; Supplemental Table S1) [21–31]. Prior to harmonization, quality control included removal of low quality cells and multiplets. Cell-type classifications for all remaining cells were obtained using the reference-based mapping pipeline Azimuth [32]. Harmonization included dimensionality reduction with the scVI method [33,34] and creation of Uniform Manifold Approximation and Projection (UMAP) plots.

### Spatial transcriptomics

We used publicly available Visium (10X Genomics) spatial transcriptomic data from the lung tissue study by Madissoon and colleagues, where they identified 80 lung cell types and states and gleaned details regarding lung micro-environments [31]. After excluding spots with low-quality data, spatial transcriptomic data from three NS bronchi samples and three CS bronchi samples were available for analyses (Supplemental Table S2). We used CytoSPACE to construct spatial datasets with high gene coverage and single-cell resolution [19], by mapping cells from the lung scRNA-seq reference atlas to spots in lung Visium transcriptomic data.

### Cell-cell interactions

CellChat (v2) enables prediction of cell-cell communication altered by smoking using data from multiple spatial transcriptomics datasets [20]. We have leveraged the capabilities of the CellChat method for use with the virtual single-cell transcriptomic data we created with CytoSPACE, where instead of RNA-seq data in each spot we have scRNA-seq data for multiple mapped cells within each spot. Likewise, the group label for each spot (typically created using a clustering method) was replaced by the cell types for each of the cells mapped from our lung scRNA-seq atlas. In this usage model, we are inferring spatially proximal cell-cell communication between interacting cell types from spatially resolved transcriptomics at virtual single-cell resolution. Additional details regarding methods are available in the Supplementary Information.

## Results

### Lung spatial transcriptomics at single-cell resolution

The lung scRNA-seq reference atlas for use with CytoSPACE was created by harmonizing data from ten prior lung tissue studies (n=104; Supplemental Table S1) [21–31]. The final harmonized dataset included 28 cell types and 349,109 cells (Supplemental Table S3). In the UMAP plot of the atlas data, we observed harmonization across all studies in the clustering by cell type (Figure 1B and Supplemental Figures S1-S11). We created lung spatial transcriptomic data at virtual single-cell resolution by mapping cells from this lung scRNA-seq atlas to spots in lung spatial transcriptomic data using CytoSPACE [19]. Scatterpie plots illustrate the cell mapping for each NS and CS sample (Figures 1C and 1D; Supplemental Figures S12-S15), where spots lacking data are either devoid of tissue or had insufficient gene expression. In these scatter plots, the epithelium was marked by higher abundances of multiciliated and smooth muscle cells. The overall number of cells mapped to the available spots in the spatial samples varies from 1 to 28, with 53% of the all spots having three or fewer cells (Supplemental Figure S16). Using hypergeometric tests, we observed enriched (p < 0.05) usage of cells from five studies for NS data and enriched usage from three studies for ever-smoker (ES) data, including Madissoon et al. (Supplemental Figures S17 and S18). Cell type availability varies across the studies (Supplemental Figures S1-S11), enabling biased usage of scRNA-seq data during cell mapping.

### Cell-cell interactions

We examined cell-cell communication altered by smoking using CellChat [20]. Spatial data for goblet cells in the NS samples were not included in the CellChat analysis, as goblet cells were not available in the ES scRNA-seq atlas data. After combining the three NS samples and the three CS samples to create two analytical datasets, and inferring the cell-cell communication network (see Supplemental Methods), we examined the differential number of interactions and interactions strength within the network between the two subjects (NS vs. CS; Supplemental Figures S19). Fibroblasts, basal and suprabasal cells and DC2 cells had higher overall differential interactions counts and interaction strength. In a scatter plot of differential signaling showing the differential communication probability for each cell type (Figure 2A), adventitial fibroblasts, basal cells and myofibroblasts are among the cells with higher incoming and outgoing signaling strength in CS with respect to NS, while DC2 cells had relatively lower overall signaling strength.

**Figure 2.**
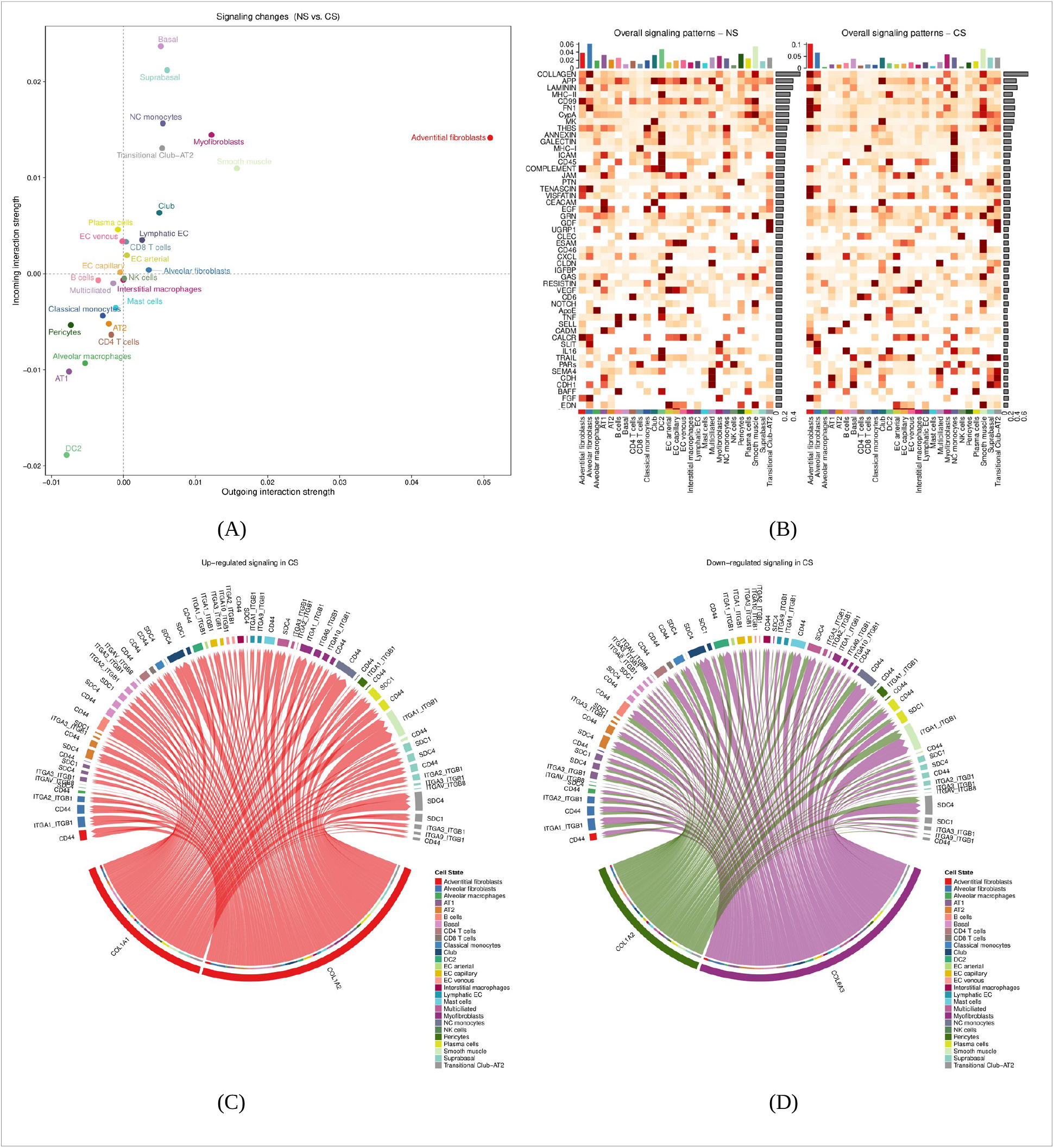
CellChat predicts cell-cell communication altered in smoker lung and highlights relevant cell types, pathways and genes. (A) scatter plot of the differential signaling (NS vs. CS) for each cell type, where the x-axis and y-axis represent the differential outgoing and incoming communication probability, (B) heatmap of the relative signaling strength of the top 40 signaling pathways across all cell types, (C) chord diagram for the top up-regulated signaling pathway (COLLAGEN), and (D) chord diagram for the top down-regulated signaling pathway (COLLAGEN).

In the ranked heatmap of overall (incoming and outgoing) relative signaling pathway strength across all cell types in the cell-cell communication network (Figure 2B), we observed collagen as the top signaling pathway. Pathway identification seeks to highlight the biological roles represented by the ligands and receptors identified in the predicted cell-cell communication network. We identified over-expressed genes by smoking condition, mapped this information on the cell-cell network, and ranked pathways by overall representation in the final network (see Supplemental Methods). Chord diagrams (Figures 2C and 2D; Supplemental Figures S20-S26) highlight the communication network cell types, ligands and potential receptor genes for the top eight up-regulated and down-regulated signaling pathways in CS samples vs. NS samples (Table 1). Some pathways (e.g. collagen) appear in both up-regulated and down-regulated columns of Table 1, as the cell types and genes involved are different. In Figures 2C and 2D, we observed increased collagen pathway activity involving the ligands COL1A1 and COL1A2 in adventitial fibroblasts and decreased activity involving COL1A2 and COL6A3 in pericytes and myofibroblasts, respectively. Receptors such as SDC4 and CD44 are predicted in several cell types, including club cells and non-classical monocytes. We also observed pathways with structural (e.g. FN1: Fibronectin-1; Fig S10), immune-related (e.g. MHC-II; Fig S11), growth factor (e.g. PTN: Pleiotrophin; Fig S10) and immunophilin (e.g. CypA: Cyclophilin A; Figure S11) roles.

**Table 1.**
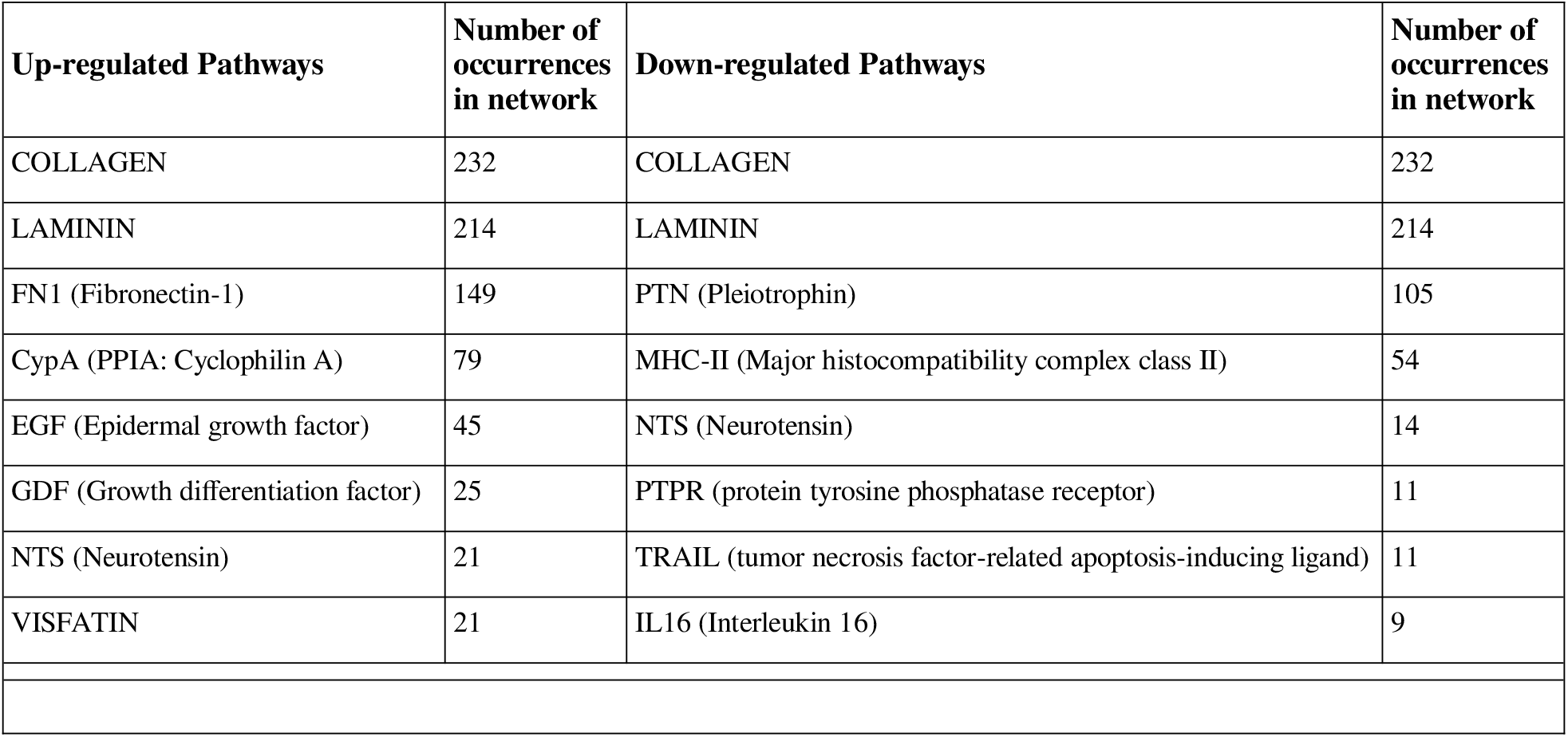
Top eight up- and down-regulated signaling pathways in cell-cell communications altered in the smoker lung.

| **Up-regulated Pathways** | **Number of occurrences in network** | **Down-regulated Pathways** | **Number of occurrences in network** |
| --- | --- | --- | --- |
| COLLAGEN | 232 | COLLAGEN | 232 |
| LAMININ | 214 | LAMININ | 214 |
| FN1 (Fibronectin-1) | 149 | PTN (Pleiotrophin) | 105 |
| CypA (PPIA: Cyclophilin A) | 79 | MHC-II (Major histocompatibility complex class II) | 54 |
| EGF (Epidermal growth factor) | 45 | NTS (Neurotensin) | 14 |
| GDF (Growth differentiation factor) | 25 | PTPR (protein tyrosine phosphatase receptor) | 11 |
| NTS (Neurotensin) | 21 | TRAIL (tumor necrosis factor-related apoptosis-inducing ligand) | 11 |
| VISFATIN | 21 | IL16 (Interleukin 16) | 9 |

## Discussion

In this study we developed a framework for analysis of spatial transcriptomics at virtual single-cell resolution. Through analyses in lung tissue samples, we identified cell types, ligands and potential receptors for several pathways altered in smoker lungs. We observed structural, inflammatory and growth factor pathways in our results, aligned with previous findings regarding molecular effects of smoking in lung disease [35–38] and current therapeutic development efforts [39]. Collagen was the top pathway identified, with adventitial fibroblasts, myofibroblasts and pericytes being the ligand-expressing cell types. In addition to its structural roles within the extracellular matrix, collagen can induce various cellular effects by acting as a ligand [40,41]. Collagen has key functions in lung tissue [42], with relevance to smoking related pathogenesis, in particular lung fibrosis [43] and COPD [44–46]. With total collagen previously found constant in smokers and increased in emphysema [47], and dynamic in matrix remodeling [48], both up-regulation and down-regulation in the collagen pathway may indicate a shift in collagen isoform and cell-type specific activity, as part of the overall remodeling process [36,49–51]. We observed up-regulation of the FN1 (fibronectin-1) pathway in CS, with myofibroblasts expressing the FN1 ligand. The fibronectin-1 pathway represents another matrix remodeling finding previously observed in smoking [52] with implications in pulmonary fibrosis [53,54], epithelial mesenchymal transition (EMT) and COPD [55,56].

The cyclophillin (CypA) pathway was up-regulated in CS samples, with the ligand PPIA (peptidylprolyl isomerase A; gene encoding CypA) expressed in myofibroblasts, lymphatic endothelial cells, suprabasal cells and basal cells. CypA is an imunophilin with molecular chaperone functions. Although typically localized in the cytoplasm, it has intercellular communication roles when secreted in response to inflammatory stimuli [57], infection [58] or oxidative stress [59]. A proteomic study found CypA up-regulated in smokers and further enhanced in smokers with COPD [60], and another study found serum levels of CypA higher in COPD with levels further elevated during acute exacerbations [61], though the particular role of CypA in lung disease is not fully understood. The predicted receptor in the CypA pathway was BSG (basigin; CD147), a gene expressed in several cells including fibroblasts and multiciliated cells. Lung expression of CD147 was found higher in smokers with COPD and may enhance mucus secretion during exposure to cigarette smoke [62]. CD147 may also have a role in smoking initiated EMT through oxidative stress signaling [63].

Limitations of this study include the small spatial transcriptomic cohort and confounding of sex with smoking condition. Our atlas also included ever-smokers, who may be either former or current smokers. The integrated spatial and single-cell transcriptomic data are from different subjects, increasing the ability to detect cell-cell communications altered in smoking by effectively expanding the study population.

Our findings further implicate several pathways previously identified, providing additional molecular context to inform future functional experiments and therapeutic avenues to mitigate pathogenic effects of smoking.

## Data Availability

The study used ONLY openly available human data that were originally located at:
Gene Expression Omnibus:
GEO: GSE143868
GEO: GSE178360
GEO: GSE136831
GEO: GSE173896
GEO: GSE171541
GEO: GSE135893
GEO: GSE227136
GEO: GSE168191
SRA: SRP318548
Zenodo: https://zenodo.org/records/8393742
https://5locationslung.cellgeni.sanger.ac.uk/
EMBL-EBI BioStudies/ArrayExpress: E-MTAB-11640

## Ethics approval and consent to participate

Data used in this study were obtained from publicly available repositories.

## Competing interests

No competing interests were disclosed for Drs. Morrow and El-Husseini

Dr. Hersh reports research grants from Bayer, Boehringer-Ingelheim and Vertex, and consulting fees from Chiesi, Sanofi and Takeda, unrelated to this study.

Dr. Yun reports research grants from Bayer, consulting fees from Bridge Biotherapeutics, and travel reimbursement from The Korean Academy of Tuberculosis and Respiratory Diseases in the past 3 years, unrelated to the current work.

## Availability of data and materials

Data were obtained from publicly available repositories (see Supplemental Tables S1 and S2):

<u>Single-cell RNA-seq data</u>

Gene Expression Omnibus

GEO: GSE143868

GEO: GSE178360

GEO: GSE136831

GEO: GSE173896

GEO: GSE171541

GEO: GSE135893

GEO: GSE227136

GEO: GSE168191

SRA: SRP318548

Zenodo: https://zenodo.org/records/8393742

https://5locationslung.cellgeni.sanger.ac.uk/

<u>Spatial Transcriptomics data</u>

EMBL-EBI BioStudies/ArrayExpress: E-MTAB-11640

## Funding

This study was supported by an Alpha-1 Foundation Research Grant, a TOPMed Fellowship and NIH grants K25HL136846, R01HL166231, P01HL114501

## Authors’ contributions

JDM: conceptualization, methodology, formal analysis, interpretation of data, manuscript preparation and approval of the final version

ZWE: methodology, interpretation of data, manuscript preparation and approval of the final version

JHY: methodology, interpretation of data, manuscript preparation and approval of the final version

CPH: methodology, interpretation of data, manuscript preparation and approval of the final version

## List of abbreviations

CS: Current-smoker
NS: Never-smoker
ES: Ever-smoker
COPD: Chronic Obstructive Pulmonary Disease
IPF: Idiopathic Pulmonary Fibrosis
scRNA-seq: Single-cell RNA-sequencing
CytoSPACE: Cellular (Cyto) Spatial Positioning Analysis via Constrained Expression alignment
UMAP: Uniform Manifold Approximation and Projection
EMT: Epithelial Mesenchymal Transition
MHC: Major Histocompatibility Complex EGF Epidermal Growth Factor
GDF: Growth Differentiation Factor
PTPR: Protein Tyrosine Phosphatase Receptor
FN1: Fibronectin-1
CypA: Cyclophilin A
NTS: Neurotensin
PTN: Pleiotrophin
MHC-II: Major histocompatibility complex class II
TRAIL: Tumor necrosis factor-related apoptosis-inducing ligand
IL16: Interleukin 16

## Notes

### Author Declarations

The study used ONLY openly available human data that were originally located at: Gene Expression Omnibus: GEO: GSE143868 GEO: GSE178360 GEO: GSE136831 GEO: GSE173896 GEO: GSE171541 GEO: GSE135893 GEO: GSE227136 GEO: GSE168191 SRA: SRP318548 Zenodo: https://zenodo.org/records/8393742 https://5locationslung.cellgeni.sanger.ac.uk/ EMBL-EBI BioStudies/ArrayExpress: E-MTAB-11640

