## Supplemental Figures S1-S11 for "Airway Spatial Transcriptomics in Smoking"

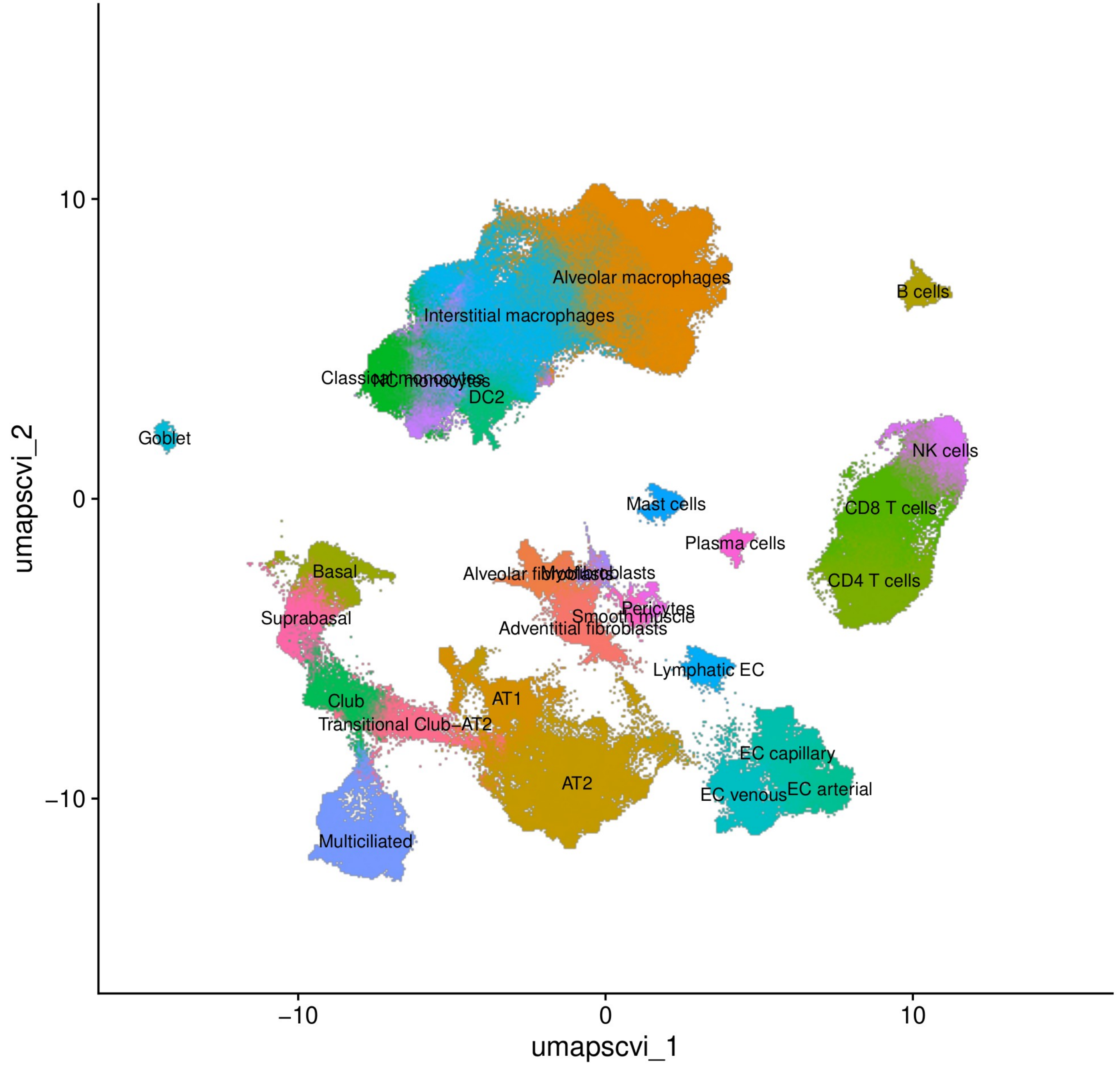


Figure S1. UMAP of scRNA-seq atlas

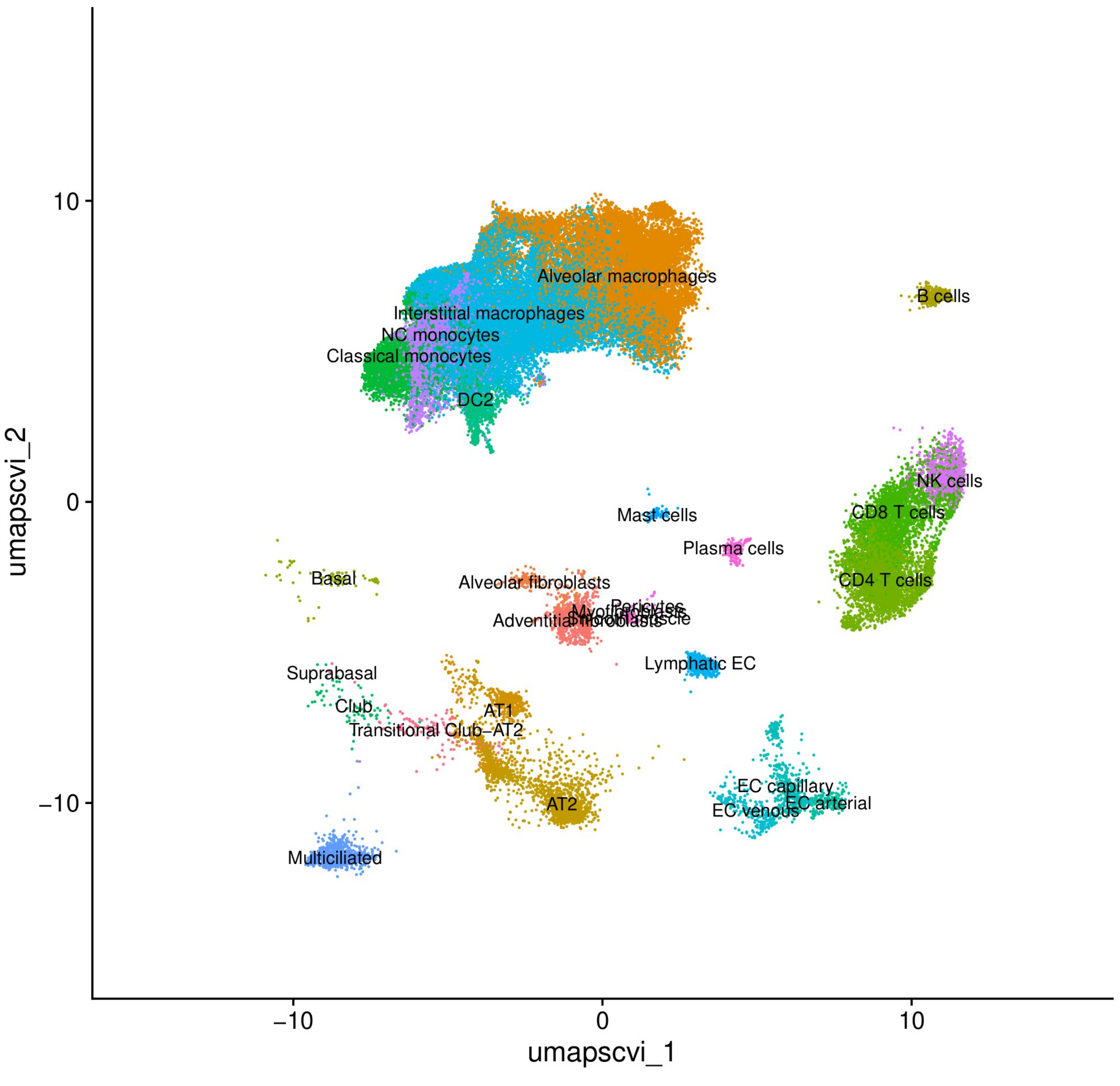


Figure S2. UMAP of Sauler et al. scRNA-seq data in atlas

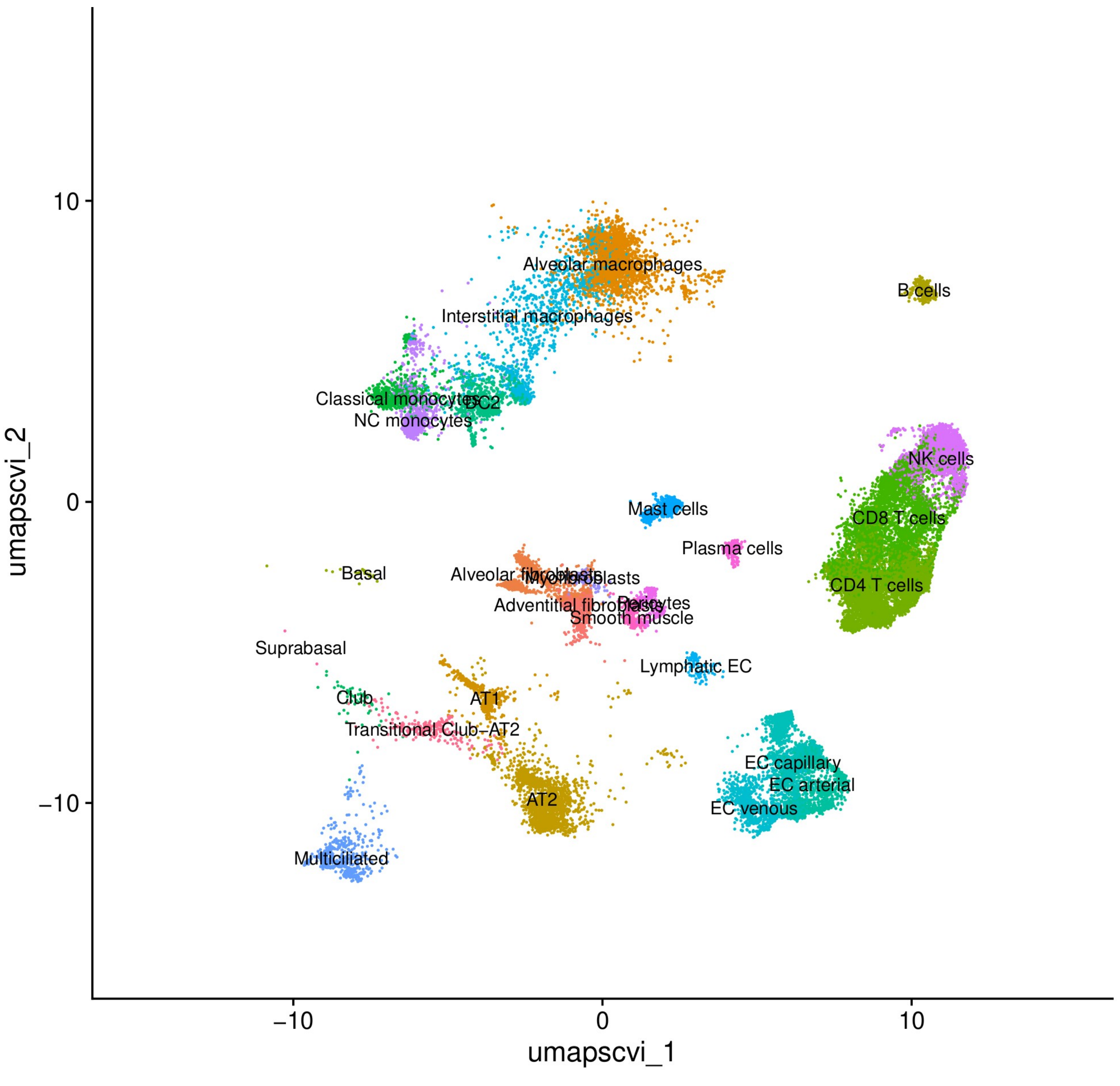


Figure S3. UMAP of Watanabe et al. scRNA-seq data in atlas

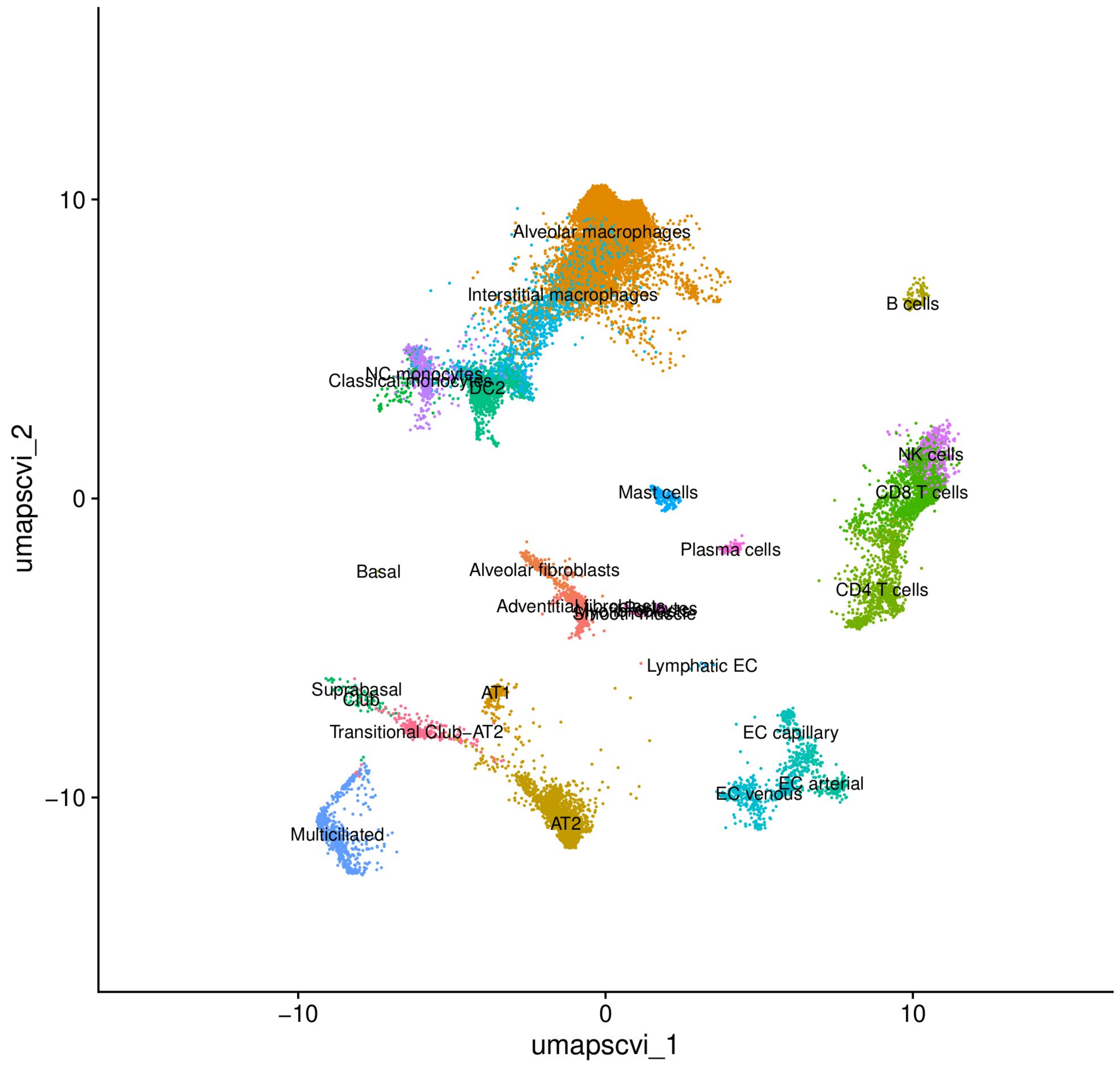


Figure S4. UMAP of Huang et al. scRNA-seq data in atlas

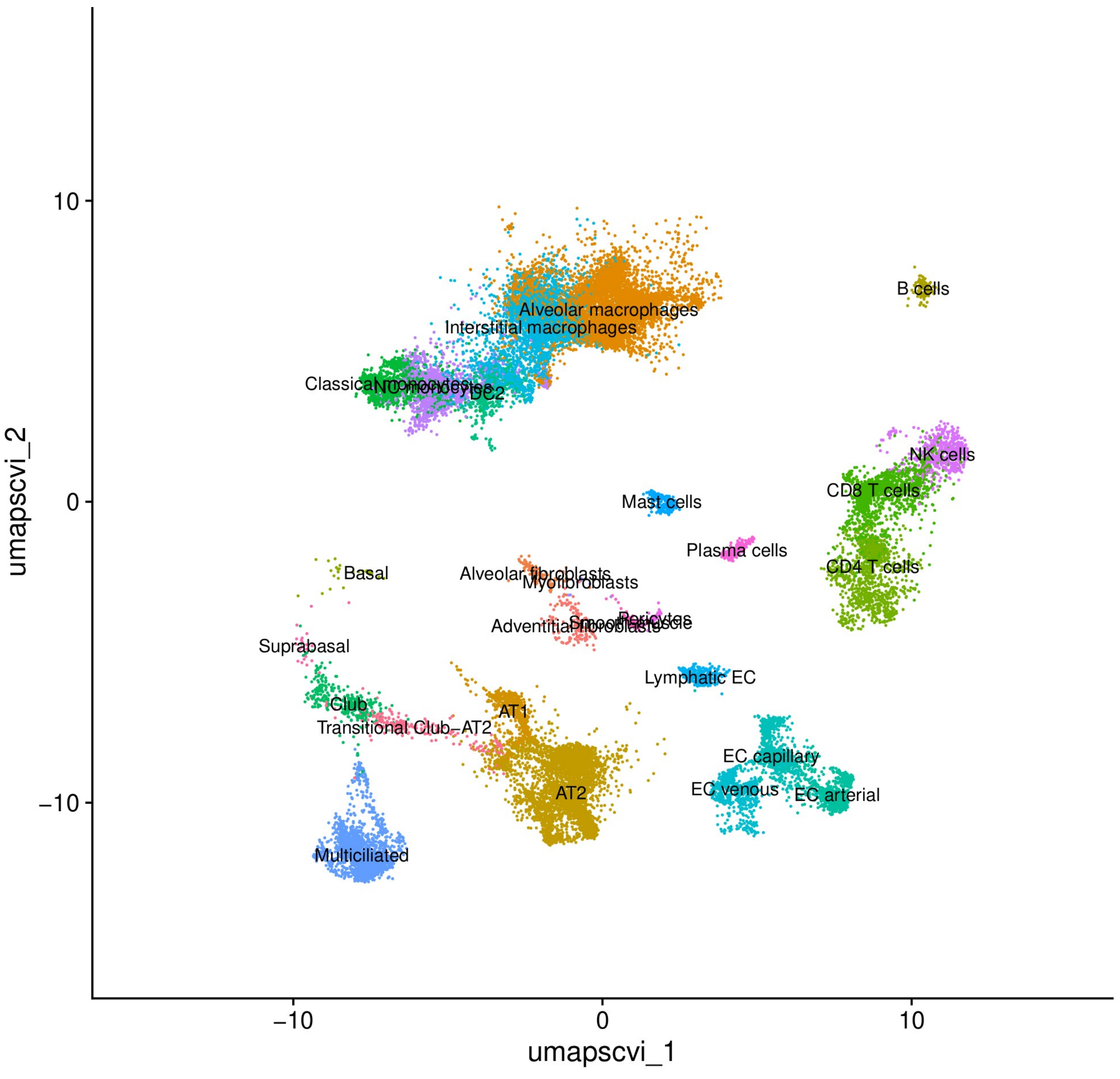


Figure S5. UMAP of Habermann et al. scRNA-seq data in atlas

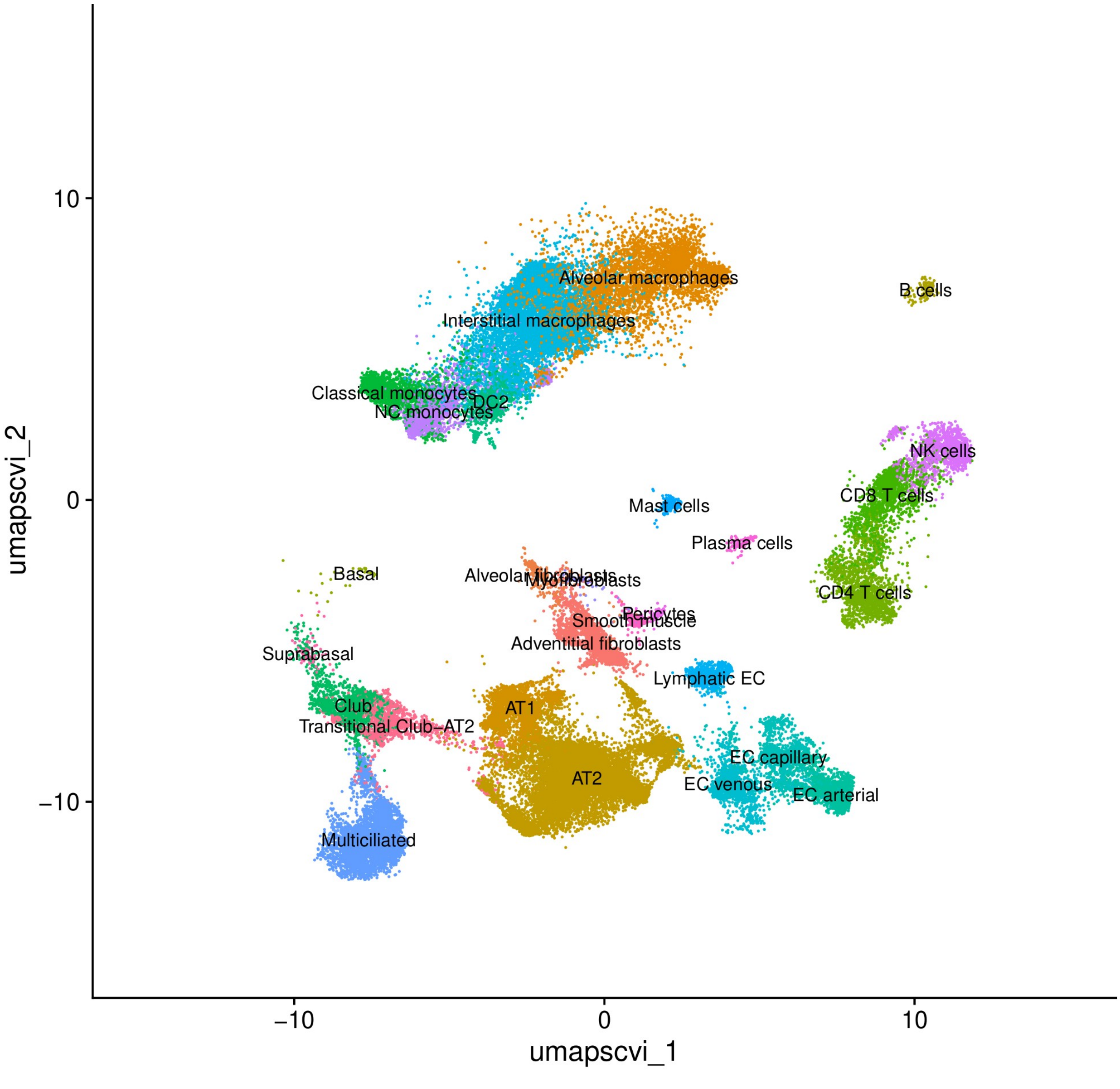


Figure S6. UMAP of Natri et al. scRNA-seq data in atlas

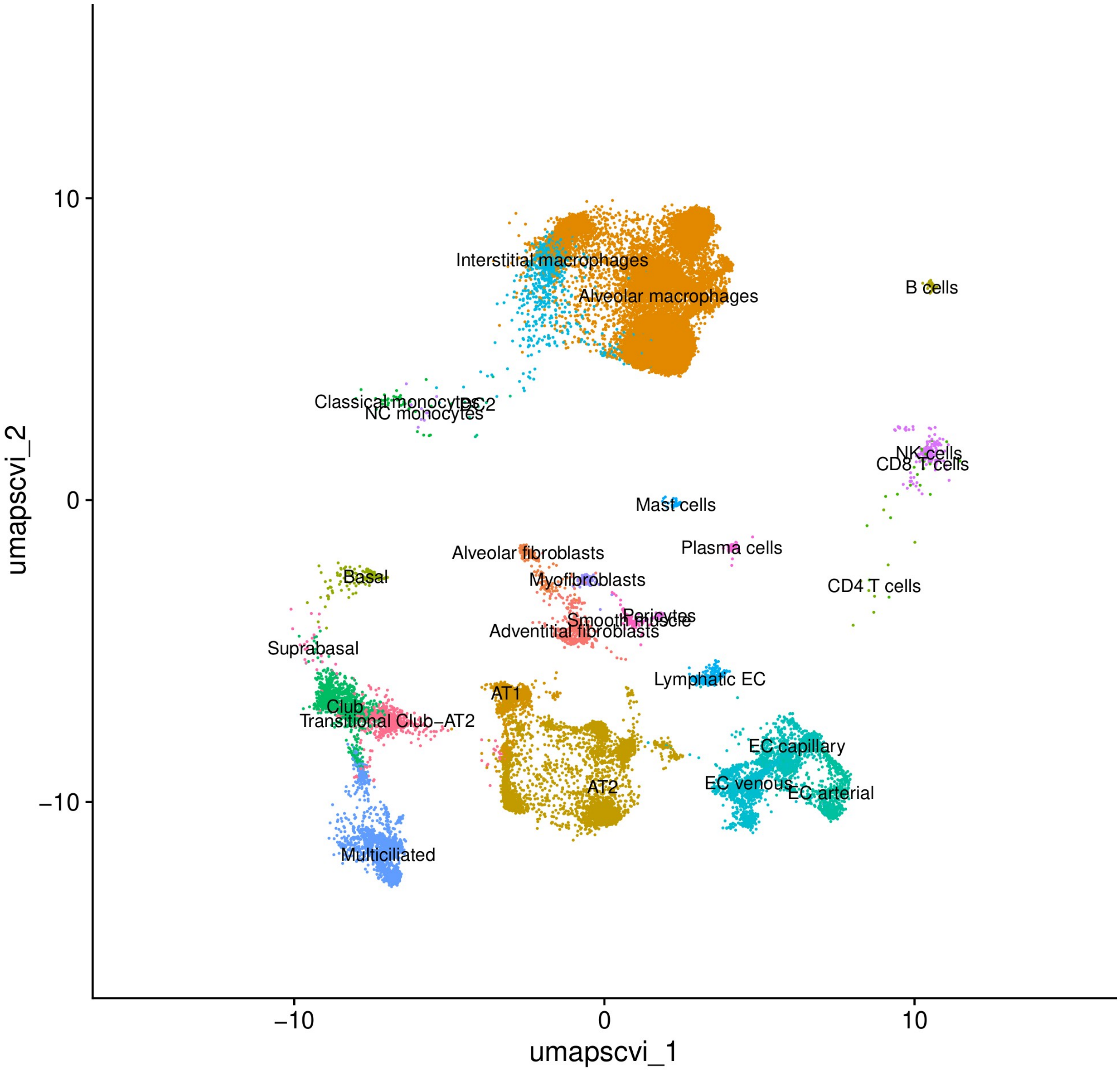


Figure S7. UMAP of Basil et al. scRNA-seq data in atlas

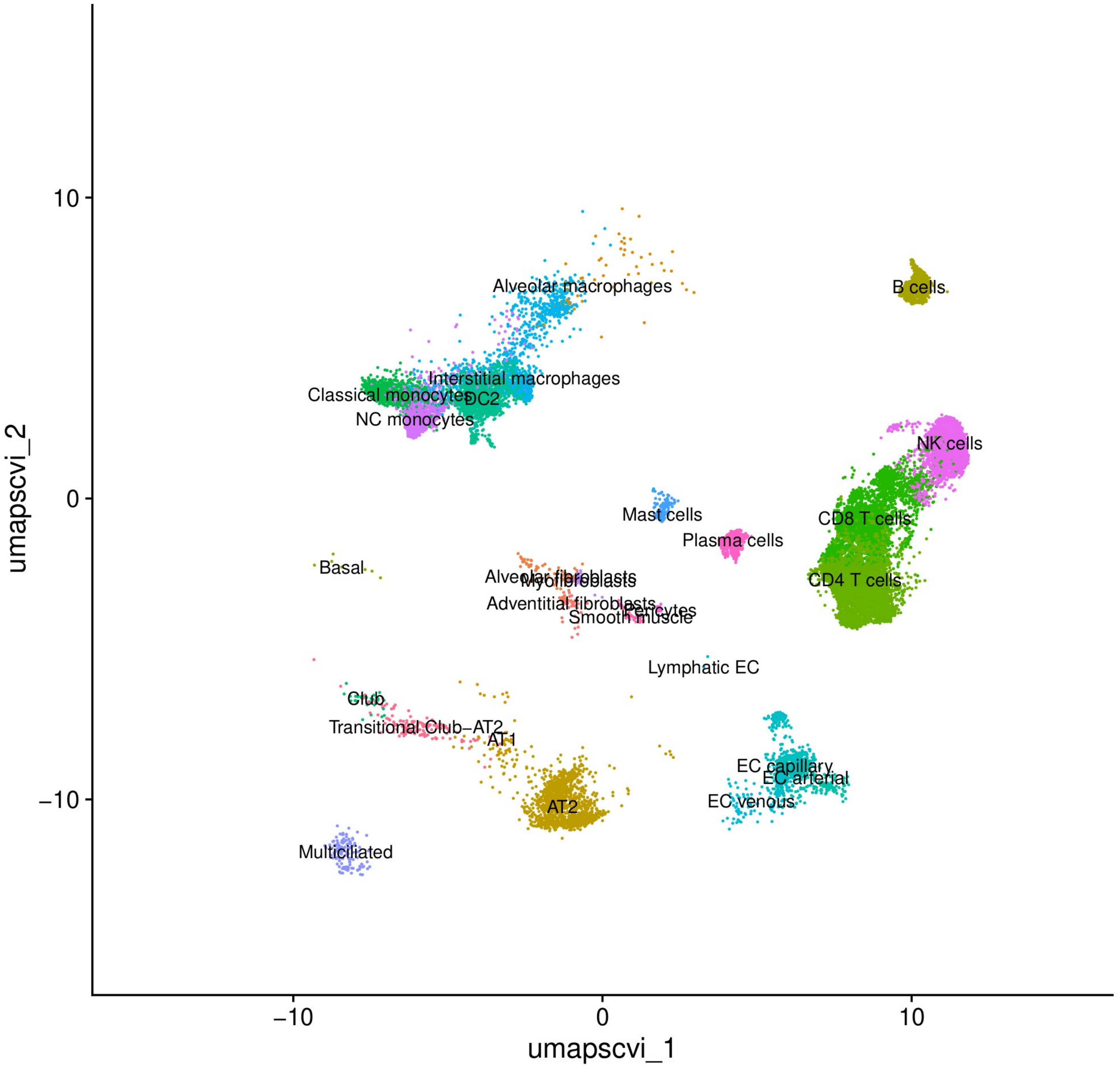


Figure S8. UMAP of Villaseñor-Altamirano et al. scRNA-seq data in atlas

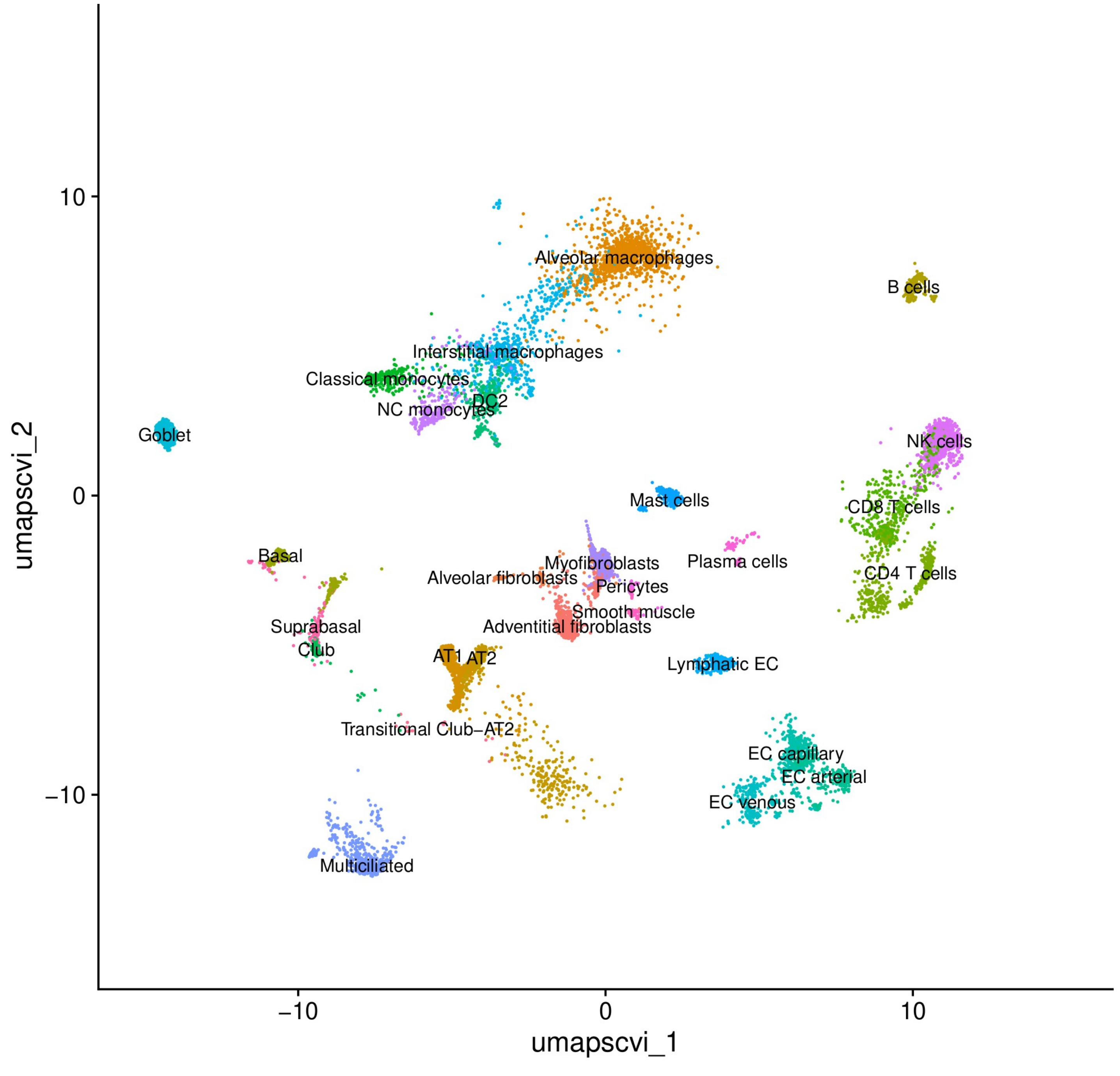


Figure S9. UMAP of Madissoon et al. scRNA-seq data in atlas

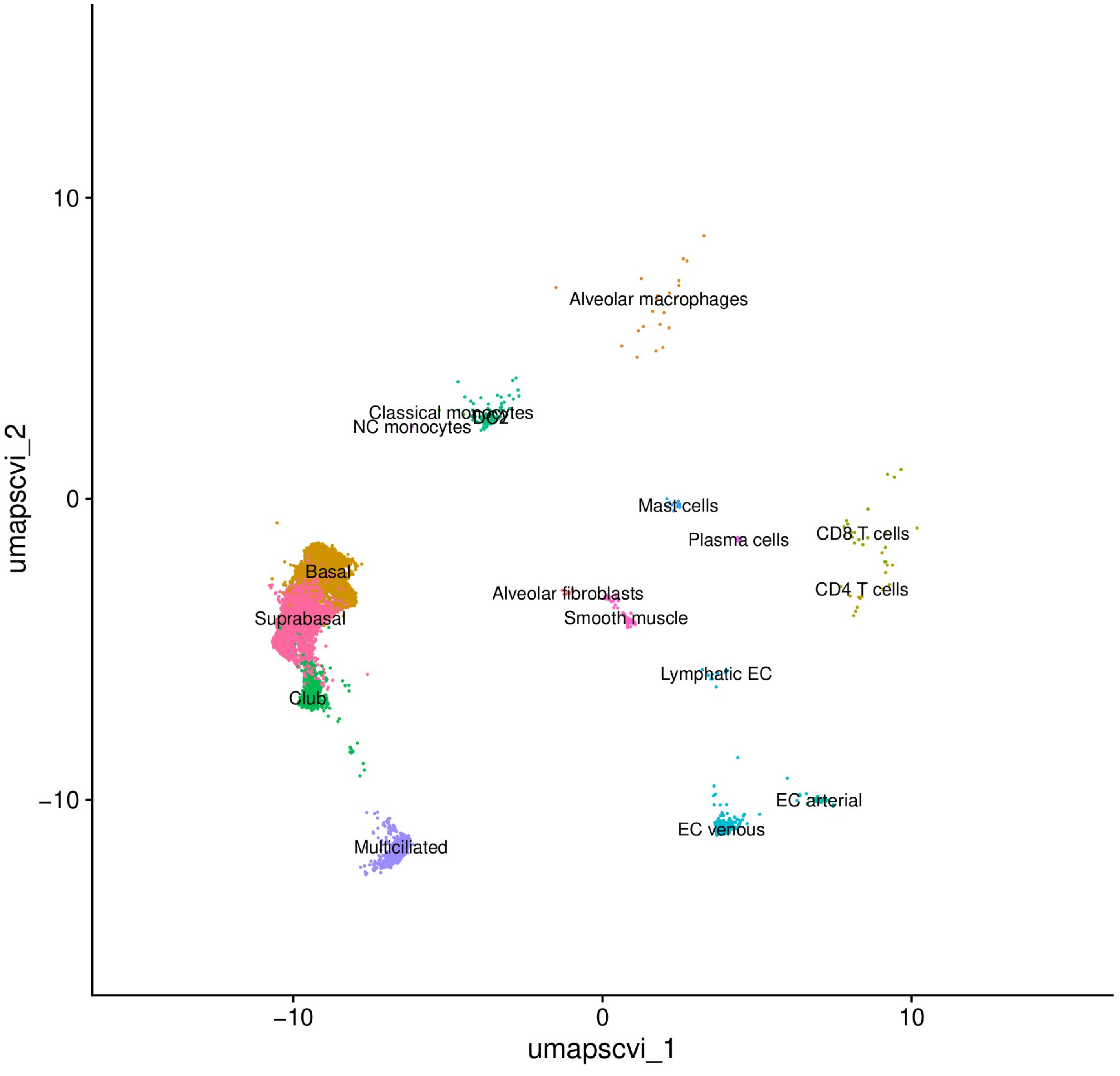


Figure S10. UMAP of Deprez et al. scRNA-seq data in atlas

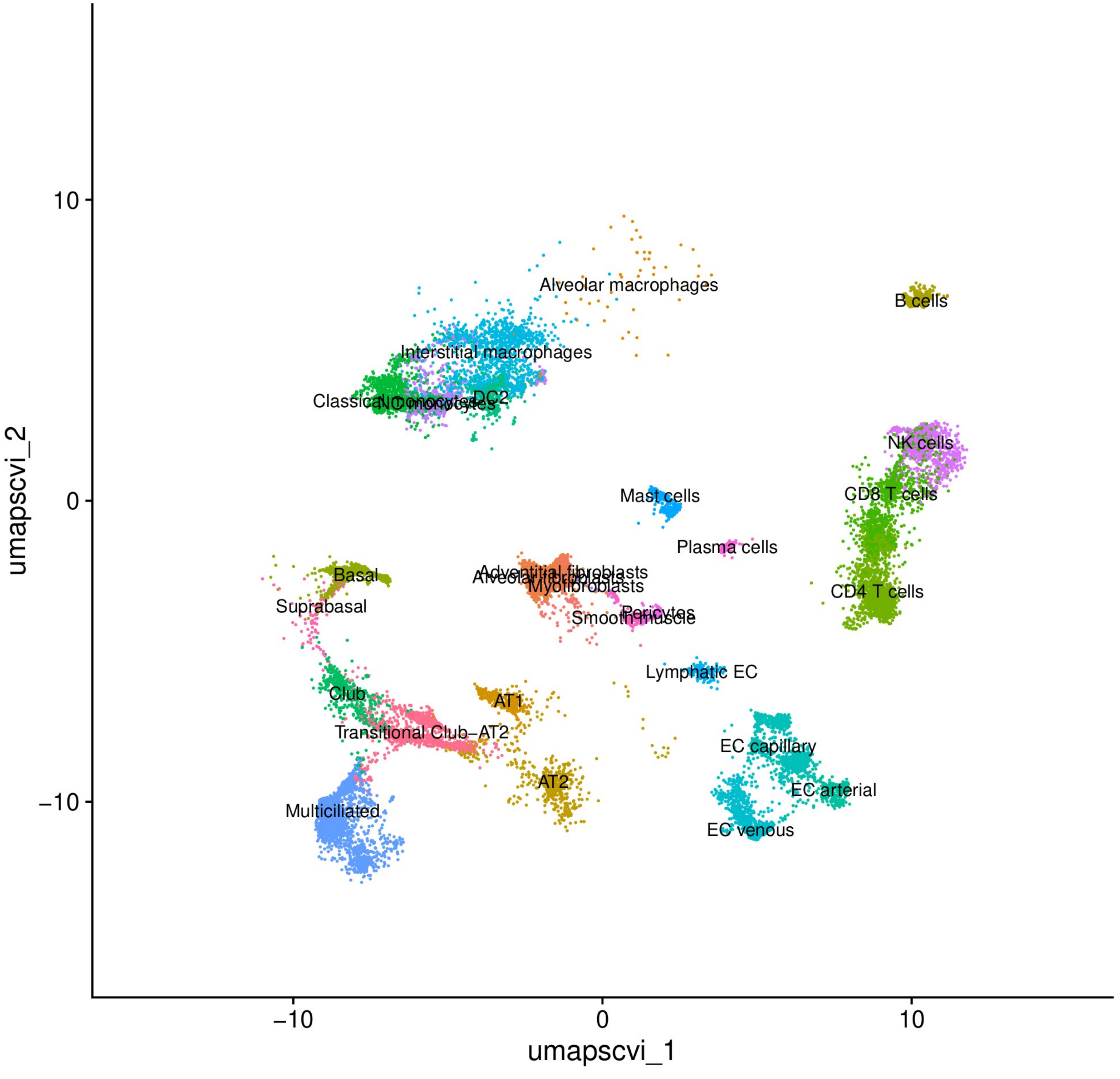


Figure S11. UMAP of Murthy et al. scRNA-seq data in atlas
