## Supplemental Material for "Airway Spatial Transcriptomics in Smoking"

**Supplemental Methods**

Lung single-cell RNA-seq data

The lung scRNA-seq reference atlas for use with CytoSPACE was created by harmonizing data from ten prior lung tissue studies (n=104; Supplemental Table S1) [1–11]. These scRNA-seq and phenotypic data for each of these studies are publicly available from the Gene Expression Omnibus (GEO), Short Read Archive (SRA), and the published articles [1–11]. Madissoon and colleagues produced single-nucleus RNA-seq data for two never-smoker and two current-smoker subjects as part of their spatial transcriptomic study of lung tissue where they identified 80 lung cell types and states, and gleaned details regarding lung micro-environments [11]. In the study of healthy human airways, Deprez and colleagues created a single-cell atlas while investigating cell population distributions and transcriptional changes along the airways [10]. Murthy et al. investigated distal airways using spatial and single-cell transcriptomics and identified unique airway cell types and trajectories with lineage and developmental relevance, while creating a distal lung map [9]. In an extensive study of cell-type specific transcriptomics and cell types in COPD, Sauler and colleagues identified novel subtypes of alveolar epithelial type II (AT2) cells relevant to COPD, among many other findings [2,8]. Watanabe and colleagues also characterized lung AT2 cells using scRNA-seq [3], and Huang et al. observed a pro-inflammatory effect of monocytes on alveolar epithelial cells in a lung study of COPD [1]. Using scRNA-seq, Habermann and colleagues examined the role of epithelial and mesenchymal cell types in pulmonary fibrosis [6], and Natri and colleagues studied the genetic control of lung gene expression in pulmonary fibrosis [7]. Basil and colleagues identified a unique secretory cell type distinct from cells found in larger proximal airways using scRNA-seq analyses [5]. A multi-omic single-cell study by Villaseñor-Altamirano et al., where particular CD8+ T-cell states in the lung were implicated in mild-moderate COPD [4].

Single-cell RNA-seq data were available from GEO for six of the subjects in the Watanabe et al. [3] dataset (Supplemental Table S1). Sequencing data for two additional subject were obtained from the SRA repository. We mapped these sequencing data to the GRCh38 human genome reference using Cell Ranger (10x Genomics) to create the scRNA-seq gene expression count matrix for all cells from these subjects. These data were concatenated with the GEO expression data to create a single dataset for the Watanabe et al. study.

Within each of the study datasets, prior to harmonization, we removed cells with greater than 15% mitochondrial expression content or less than 1,000 transcripts. Cells identified as multiplets using Scrublet [12] were also excluded. Cell-type classifications for all remaining cells were obtained using the reference-based mapping pipeline Azimuth [13]. The Azimuth pipeline [13] matched expression profiles of each cell in the scRNA-seq data with specific cell types annotated in the scRNA-seq data from the Human Lung Cell Atlas [14]. After concatenating the annotated data from the ten studies (merging of ten Seurat objects in R), cell types with a mean cell count per subject less than ten were removed. Endothelial Cell (EC) general capillary and aerocyte capillary cells were grouped as EC capillary cells and EC venous pulmonary and EC venous systemic cells were grouped as EC venous cells. Lymphatic EC mature cells were relabeled as Lymphatic EC cells and Basal resting cells were relabeled as Basal. We exported the merged data in adata format and used the scVI method within the Python single-cell variational inference tools (scvi-tools) package [15,16] to perform dimensionality reduction of the merged expression data. We created a Uniform Manifold Approximation and Projection (UMAP) using the scVI output to identify outlying cells not within cell-type clusters. A cluster was defined by an ellipse centered on the UMAP values for a particular cell type. The ellipse size was 2*MAD (median absolute deviation) along each axis of the ellipse. Cells outside each of their respective cell-type cluster ellipse were excluded from later analyses. A final UMAP plot was used to observe harmonization of the datasets.

Spatial transcriptomics

In this study, we used the publicly available Visium (10X Genomics) spatial transcriptomic data from the lung tissue study by Madissoon and colleagues [11]. A Seurat object was created using the spatial data for each sample using the R package Seurat [13]. Spots within the Visium data with fewer than 500 counts in the gene expression data were excluded from the analyses. Although data were available from lung parenchyma samples, this low-expression filtering step yielded too few spots for analysis in the data for these sample. Spatial transcriptomic data from three never-smoker bronchi samples and three current-smoker bronchi samples were available for analyses following this filtering (Supplemental Table S2). Within each sample, genes with zero counts across all spots were excluded from the dataset.

Spatial transcriptomic data at single-cell resolution

Integrating Spatial and Single-cell Transcriptomic data leverages the strength of both technologies to provide a spatially resolved perspective on tissue cell-cell communications at single-cell resolution. We used the Python package CytoSPACE to construct spatial datasets with high gene coverage and spatially-resolved scRNA-seq data [17], by mapping cells from the lung scRNA-seq reference atlas to spots in lung Visium transcriptomic data. For the current-smoker (CS) Visum samples, we used the cells within the scRNA-seq atlas from the ever-smoker subjects. Likewise, for the never-smoker (NS) samples, we used scRNA-seq data from the never-smokers. We downsampled the scRNA-eq data to create an atlas containing no more than 1,000 cells for each cell type. Only genes represented in both the scRNA-seq atlas and the spatial sample were included in the data used in the CytoSPACE mapping process. The cells mapped to each spot in the spatial transcriptomic output data are located at the center of their assigned spot. The mapped cell type proportions across the Visium spots for each sample were viewed using the plotSpatialScatterpie function in the R package SPOTlight [18], where use of color by cell type was made consistent across all samples. We examined the usage of cells during the mapping process with respect to study source using bar plots for both CS and NS samples, and tested for enrichment of particular study data using hypergeometric tests, after adjusting for the downsampling of atlas cells prior to CytoSPACE mapping. During quality assessment, we examined the mapped cell count distribution across all spots for both the NS and CS samples.

Cell-cell interactions

CellChat (v2) enables prediction of differential cell-cell communication by smoking using data from multiple spatial transcriptomics datasets [19]. The method was developed for use with spatial transcriptomics in a format similar to the Visium (10x Genomics) platform, with RNA-seq data stored for each spot within a grid that covers the assayed tissue area. We have leveraged the capabilities of the CellChat method for use with the virtual single-cell transcriptomic data we created with CytoSPACE. Instead of a single set of RNA-seq data in each spot, our approach has scRNA-seq data for multiple mapped cells within each spot, with the mapped cells located at the center of their assigned spot. Likewise, the group label for each spot (typically created using a clustering method) was replaced by the cell types for each of the mapped cells from our lung scRNA-seq atlas. In this usage model, we are inferring spatially proximal cell-cell communication between interacting cell types from spatially resolved transcriptomics at virtual single-cell resolution.

In the analyses we used the CellChat human database, specifically the subset for only Cell-Cell Contact, ECM-Receptor, and Secreted Signaling communication. The three samples for each subject (CS and NS; Supplemental Table S2) were combined into one CellChat object to create two analytical output datasets for observation of smoking effects (CS vs. NS) on cell-cell communication. We inferred the cell-cell communication network in each subject using the computeCommunProb function in CellChat [19] with the triMean method for computing the average gene expression per cell group, an interaction range of 250 micrometers, and no distance constraints to compute communication probability. We then inferred the probability at the signaling pathway level using the CellChat function computeCommunProbPathway. Heatmaps were created using the netVisual_heatmap function to illustrate the differential number of interactions and interaction strength in the cell-cell communication network between the two subjects (NS vs. CS). Also with a focus on cell types, we created scatter plots of differential signaling using the CellChat function netAnalysis_diff_signalingRole_scatter, where the x-axis and y-axis represent the differential outgoing and incoming communication probability for each cell type and positive or negative values correspond to an increase or decrease in CS, respectively. To examine pathway relevance, we used the CellChat function netAnalysis_signalingRole_heatmap to create a heatmap of the relative signaling strength of the top signaling pathways across all cell types.

Seeking additional rigor and context for the impact of smoking on cell-cell communication, we identified signaling genes over-expressed by smoking condition for each cell type using the CellChat function identifyOverExpressedGenes, with a fold change threshold of 1.5 and p-value cutoff of 0.5/27 (27 being the number of cell types tested in the analysis). The netMappingDEG function mapped the differential expressed genes onto the cell-cell communication results, where only differential expression of the ligands (senders) was considered in the analyses. After filtering for up- and down-regulated ligands meeting the threshold cutoffs for p-value and fold change, the pathways of interest were the ones most represented among the final cell-cell communication findings. We created chord diagrams using the CellChat function netVisual_chord_gene to visually examine the differential cell-cell communication (cell types and genes) in the signaling pathways of interest. This function was modified to produce consistent color assignment for each cell type across chord diagrams for all pathways.

#### Supplemental Tables

Table S1. Single-cell RNA-seq data sources with subject demographics

| ≈ 350,000 cells | Smoking Status (n=104) | | | |
|---|---|---|---|---|
| **Data Source *** | **Never-smokers** | **Ever-smokers** | **Female / Male** | **Repository** |
| Madissoon | 2 | 2 | 2 / 2 | https://5locationslung.cellgeni.sanger.ac.uk/ |
| Deprez | 10 | 0 | 6 / 4 | GEO: GSE143868 |
| Murthy | 1 | 2 | 0 / 3 | GEO: GSE178360 |
| Sauler | 22 | 6 | 12 / 16 | GEO: GSE136831 |
| Watanabe | 3 | 5 | 3 / 5 | GEO: GSE173896 (6 samples)<br>SRA: SRP318548 (2 samples) |
| Huang | 4 | 2 | 0 / 6 | GEO: GSE171541 |
| Habermann # | 2 | 8 | 3 / 7 | GEO: GSE135893 |
| Natri # | 12 | 12 | 7 / 17 | GEO: GSE227136 |
| Basil | 3 | 2 | 3 / 2 | GEO: GSE168191 |
| Villaseñor-Altamirano | 4 | 2 | 5 / 1 | https://zenodo.org/records/8393742 |
| **Total Subjects** | **63** | **41** | **41 / 63** | |

* First author of primary study article
### Control subjects only

Table S2. Summary of spatial transcriptomic samples with subject demographics

| Subject ID | Smoking Status | Sex | Age (years) | Sample name* | Location |
|---|---|---|---|---|---|
| A42 | Never-smoker | Male | 60-64 | WSA_LngSP8759311 | Bronchi at the fourth generation |
| | | | | WSA_LngSP8759312 | |
| | | | | WSA_LngSP8759313 | |
| A37 | Current-smoker | Female | 55-60 | WSA_LngSP9258468 | Bronchi at the second/third generation |
| | | | | WSA_LngSP10193348 | |
| | | | | WSA_LngSP9258464 | |

* EMBL-EBI BioStudies/ArrayExpress: E-MTAB-11640

Table S3. Cell type distribution in the lung scRNA-seq atlas

| **Cell type** | **Ever smoker cell counts** | **Never smoker cell counts** | **Cell counts for all subjects** |
|---|---:|---:|---:|
| Alveolar macrophages | 27637 | 51321 | 78958 |
| Interstitial macrophages | 20744 | 27923 | 48667 |
| AT2 | 17305 | 18820 | 36125 |
| CD4 T cells | 10314 | 11483 | 21797 |
| CD8 T cells | 8312 | 10702 | 19014 |
| Multiciliated | 6586 | 9627 | 16213 |
| Classical monocytes | 5933 | 7310 | 13243 |
| NK cells | 5314 | 7489 | 12803 |
| NC monocytes | 4116 | 7980 | 12096 |
| EC capillary | 5472 | 4419 | 9891 |
| AT1 | 5215 | 3715 | 8930 |
| Basal | 1024 | 7448 | 8472 |
| EC venous | 3799 | 4222 | 8021 |
| DC2 | 2853 | 4447 | 7300 |
| EC arterial | 3021 | 3007 | 6028 |
| Club | 1992 | 3815 | 5807 |
| Adventitial fibroblasts | 2045 | 3518 | 5563 |
| Transitional Club-AT2 | 1707 | 3335 | 5042 |
| Suprabasal | 273 | 4586 | 4859 |
| Alveolar fibroblasts | 2586 | 1531 | 4117 |
| B cells | 1112 | 2757 | 3869 |
| Lymphatic EC | 1823 | 1706 | 3529 |
| Mast cells | 1822 | 978 | 2800 |
| Plasma cells | 637 | 1184 | 1821 |
| Goblet | 0 | 1088 | 1088 |
| Smooth muscle | 653 | 423 | 1076 |
| Myofibroblasts | 197 | 798 | 995 |
| Pericytes | 625 | 360 | 985 |
| **Total Cells** | **143117** | **205992** | **349109** |

**Supplemental Figures**

Figures S1-S11. UMAP plots of scRNA-seq atlas for all studies, and Sauler et al., Watanabe et al., Huang et al., Habermann et al., Natri et al., Basil et al., Villaseñor-Altamirano et al., Madissoon et al., Deprez et al., Murthy et al. subsets

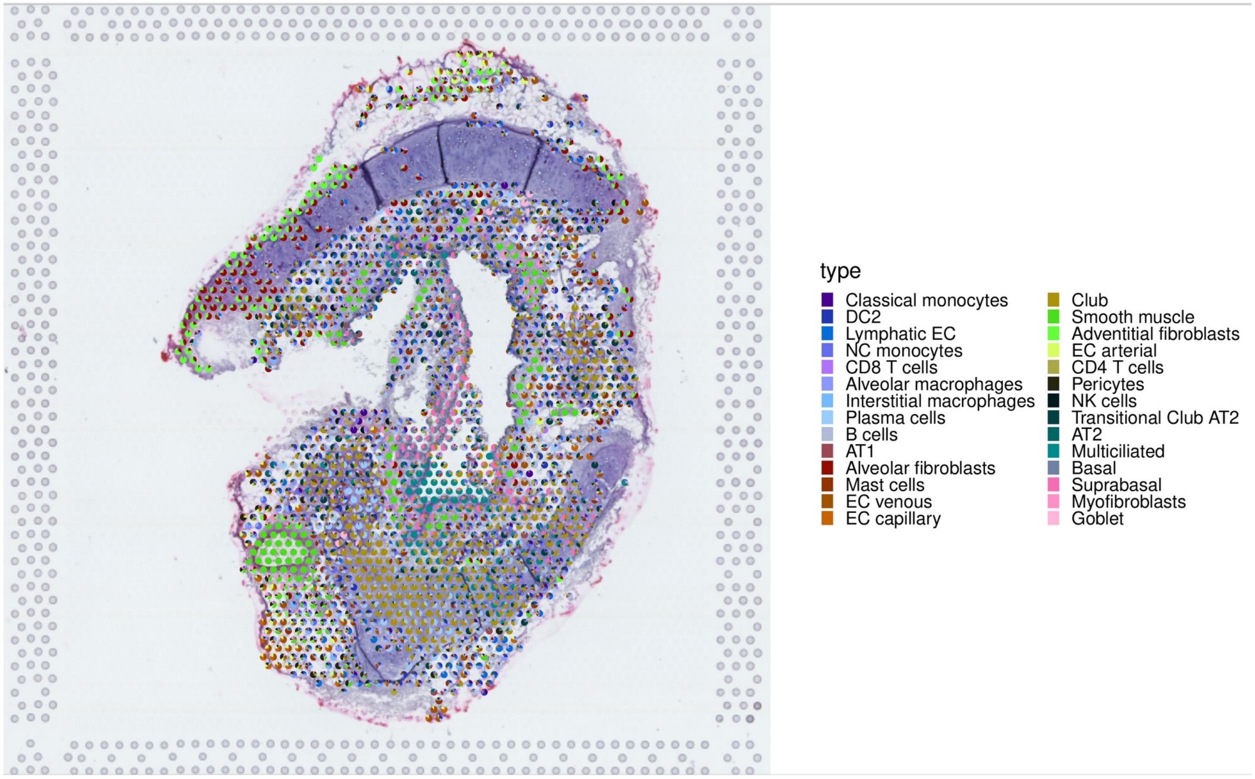


Figure S12. The cell-type composition for never-smoker sample WSA_LngSP8759311 illustrated in a scatterpie plot.

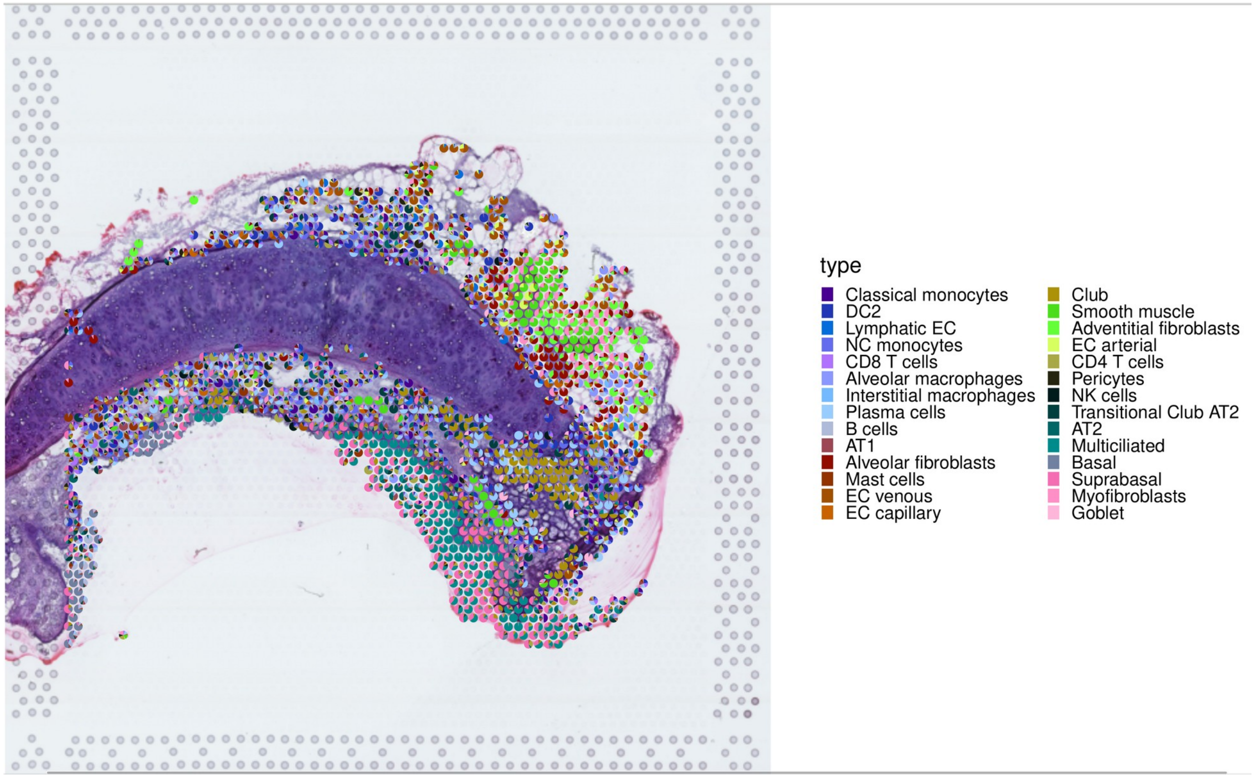


Figure S13. The cell-type composition for never-smoker sample WSA_LngSP8759313 illustrated in a scatterpie plot.

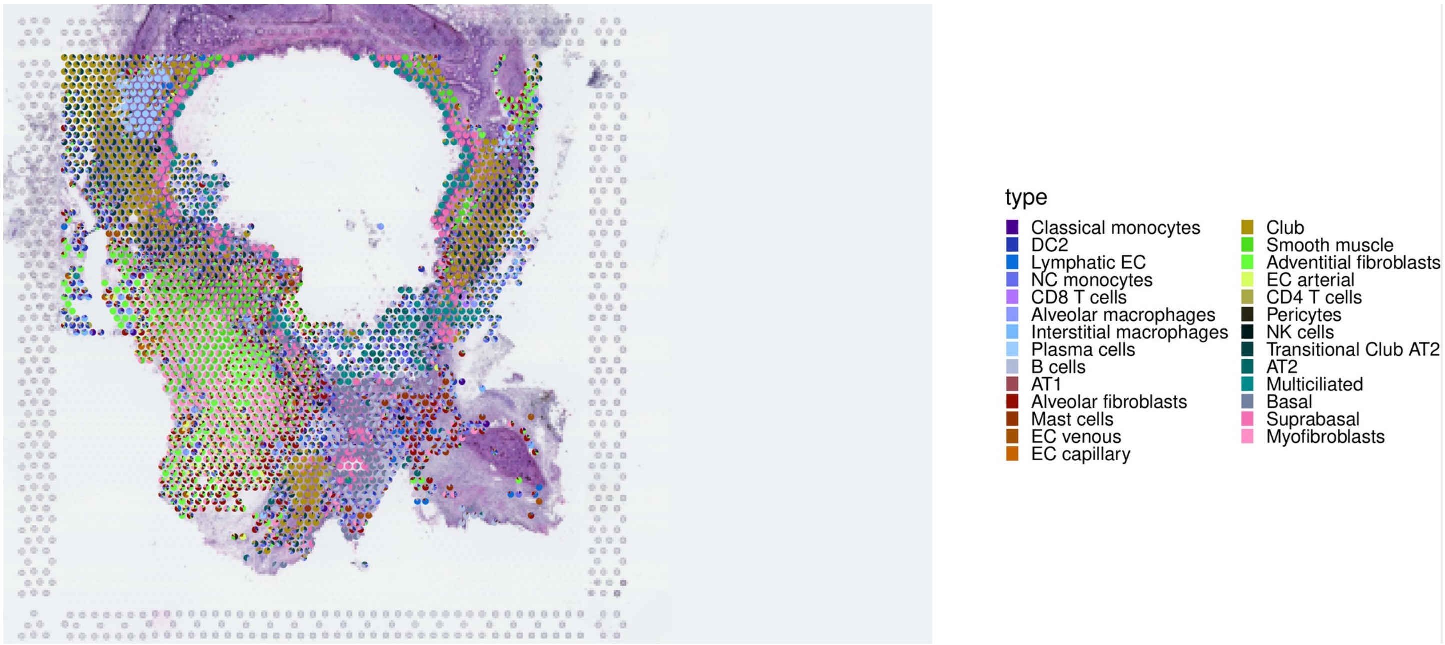


Figure S14. The cell-type composition for current-smoker sample WSA_LngSP9258468 illustrated in a scatterpie plot.

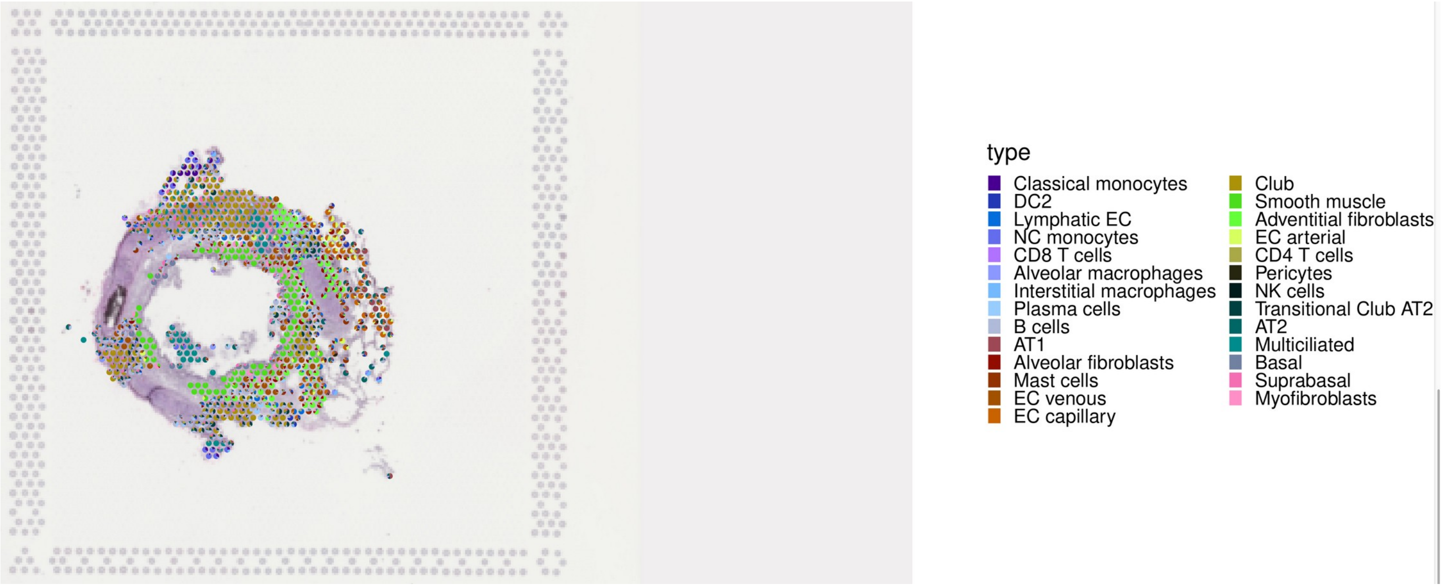


Figure S15. The cell-type composition for current-smoker sample WSA_LngSP10193348 illustrated in a scatterpie plot.

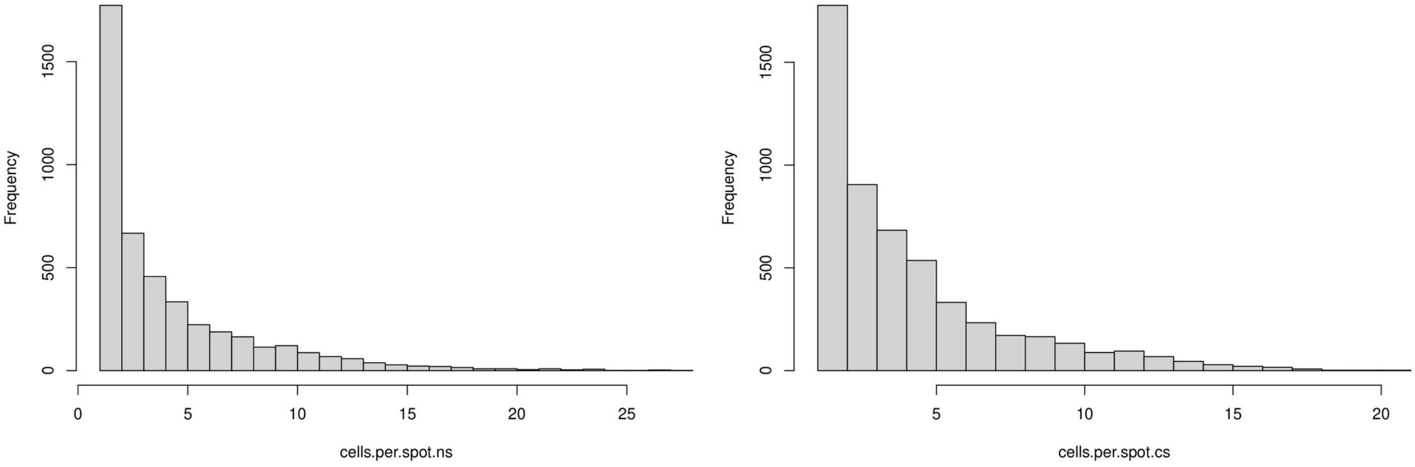


(A) (B)

Figure S16. Histogram of the number of cells mapped to spots in the spatial data for (A) never-smoker samples and (B) current-smoker samples.

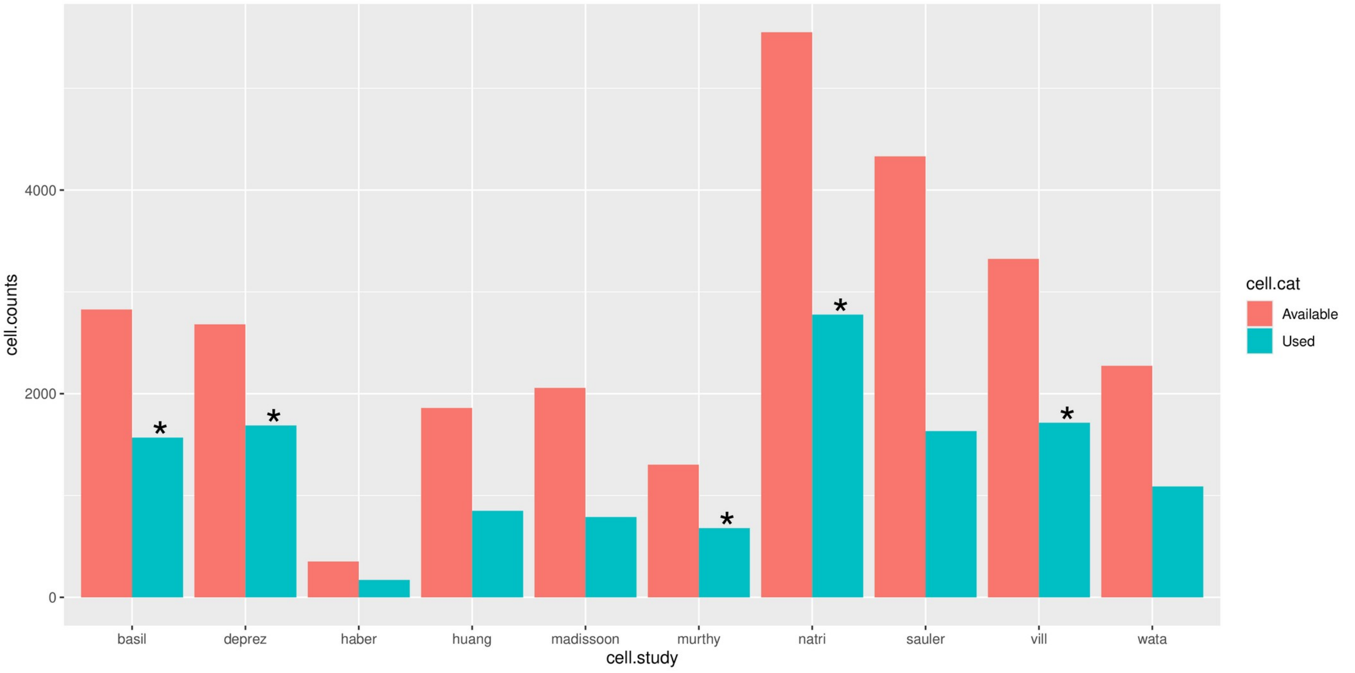


Figure S17. Bar plot of cell usage by CytoSPACE for the never-smoker scRNA-seq atlas data. Significant enrichments ($p < 0.05$) are labeled with an asterisk.

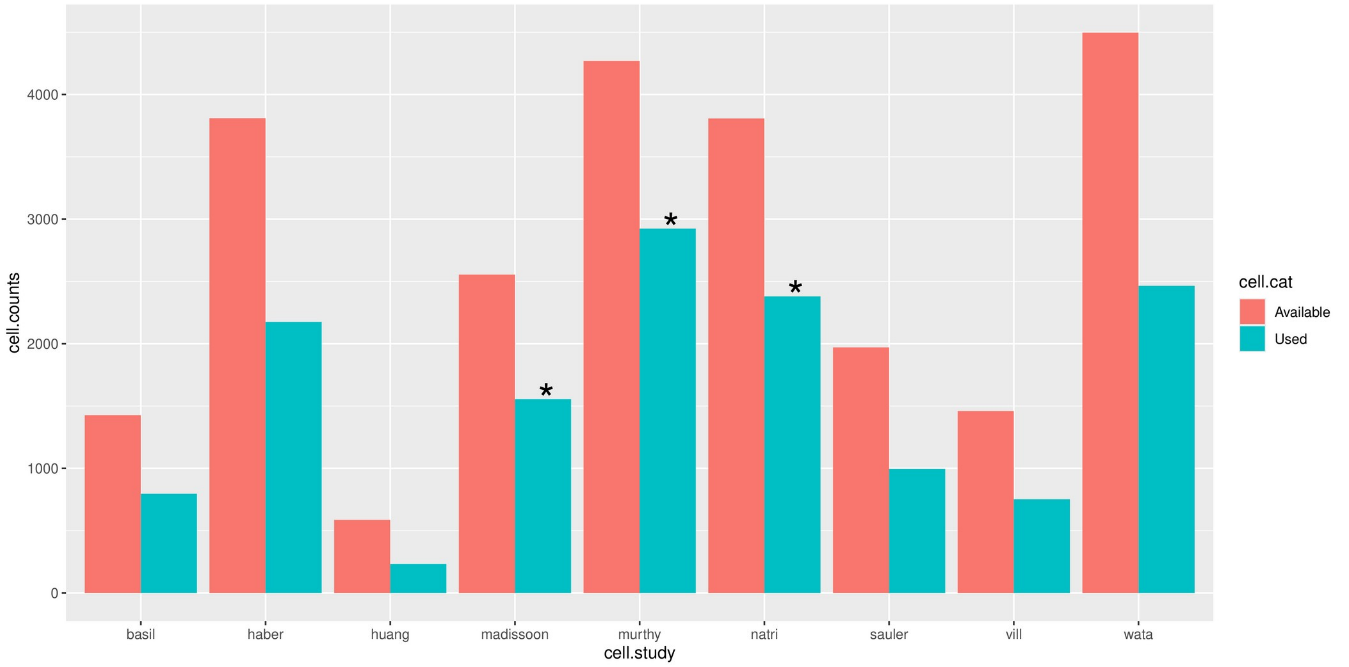


Figure S18. Bar plot of cell usage by CytoSPACE for the ever-smoker scRNA-seq atlas data. Significant enrichments ($p < 0.05$) are labeled with an asterisk.

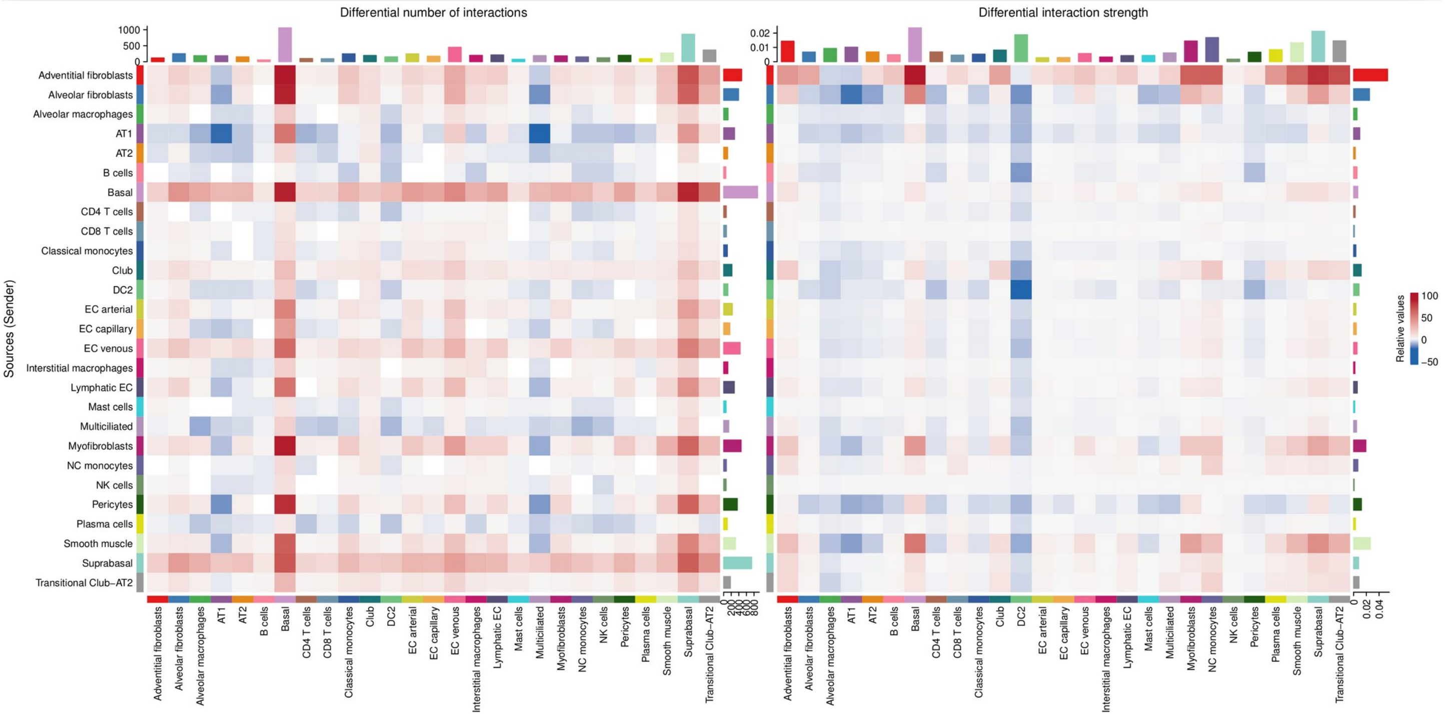


(A) (B)

Figure S19. Heatmaps of the (A) differential number of interactions and (B) the differential interaction strength within the CellChat cell-cell communication network between the two subjects (NS subject vs. CS subject)

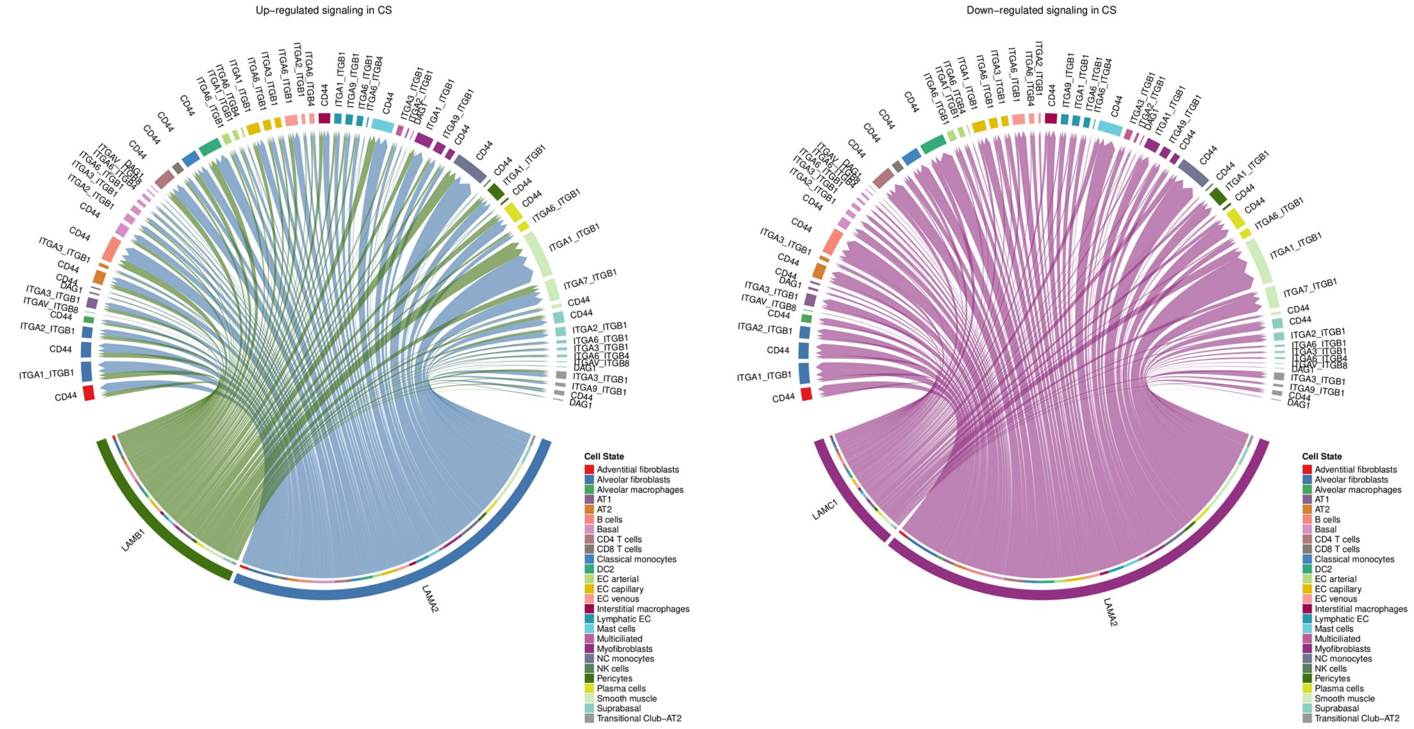


Figure S20. Chord diagrams for (A) the up-regulated signaling pathway LAMININ, and (B) the down-regulated signaling pathway LAMININ.

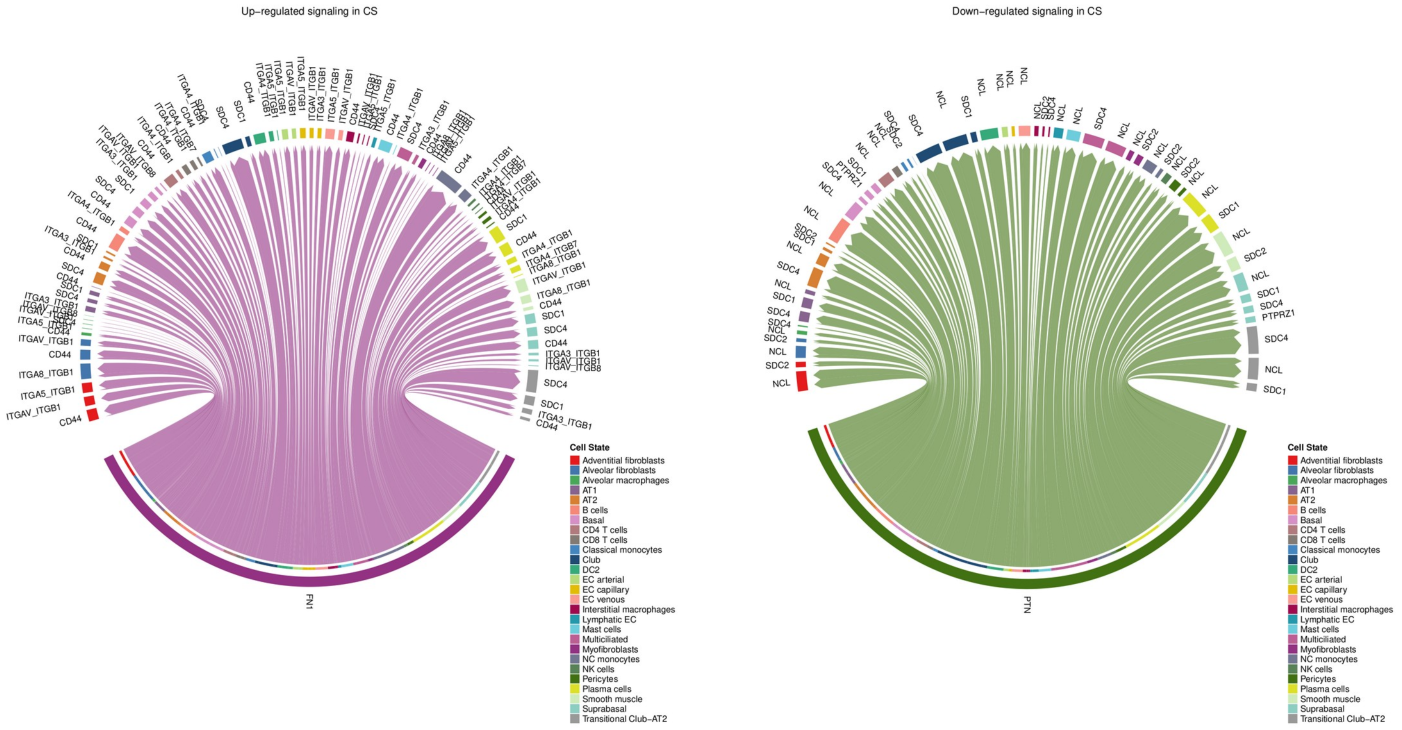


Figure S21. Chord diagrams for (A) the up-regulated signaling pathway FN1 (Fibronectin-1), and (B) the down-regulated signaling pathway PTN (Pleiotrophin).

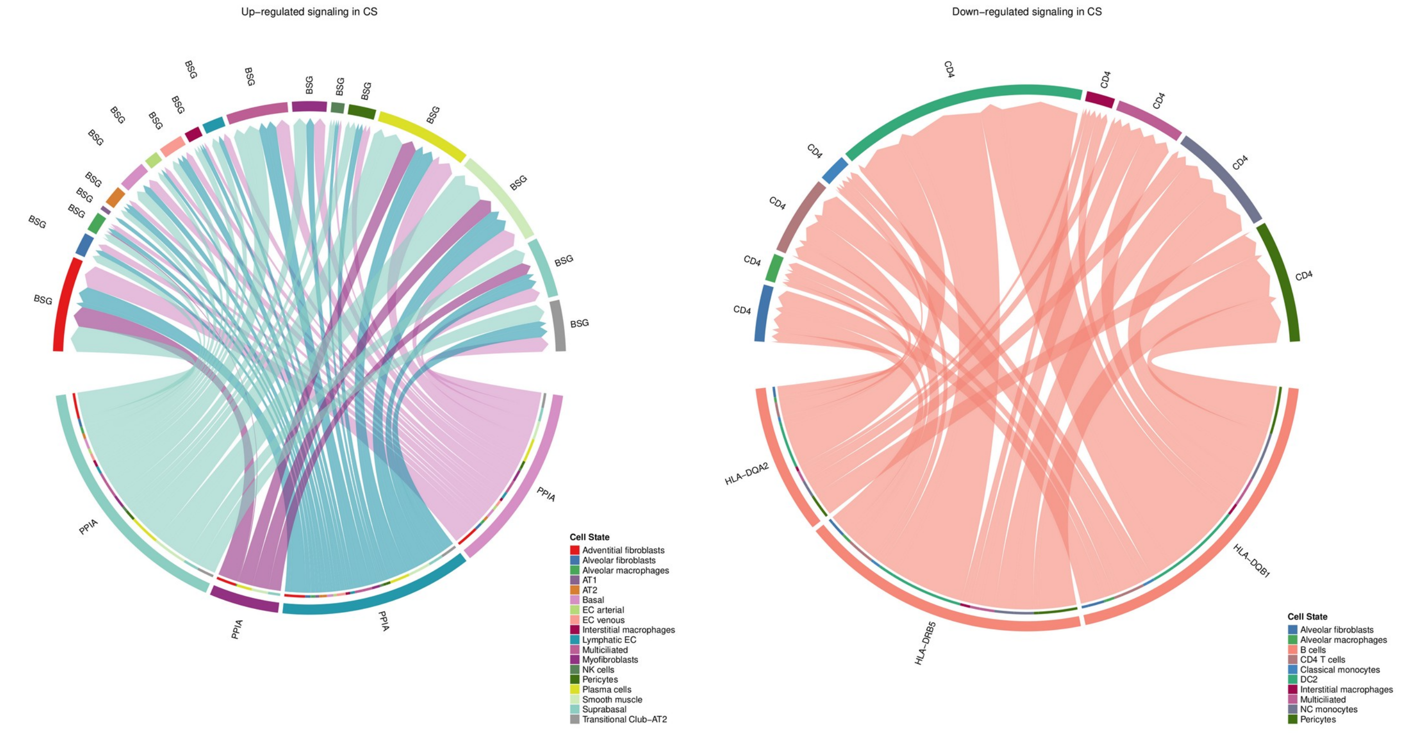


Figure S22. Chord diagrams for (A) the up-regulated signaling pathway CypA (PPIA: Cyclophilin A), and (B) the down-regulated signaling pathway MHC-II.

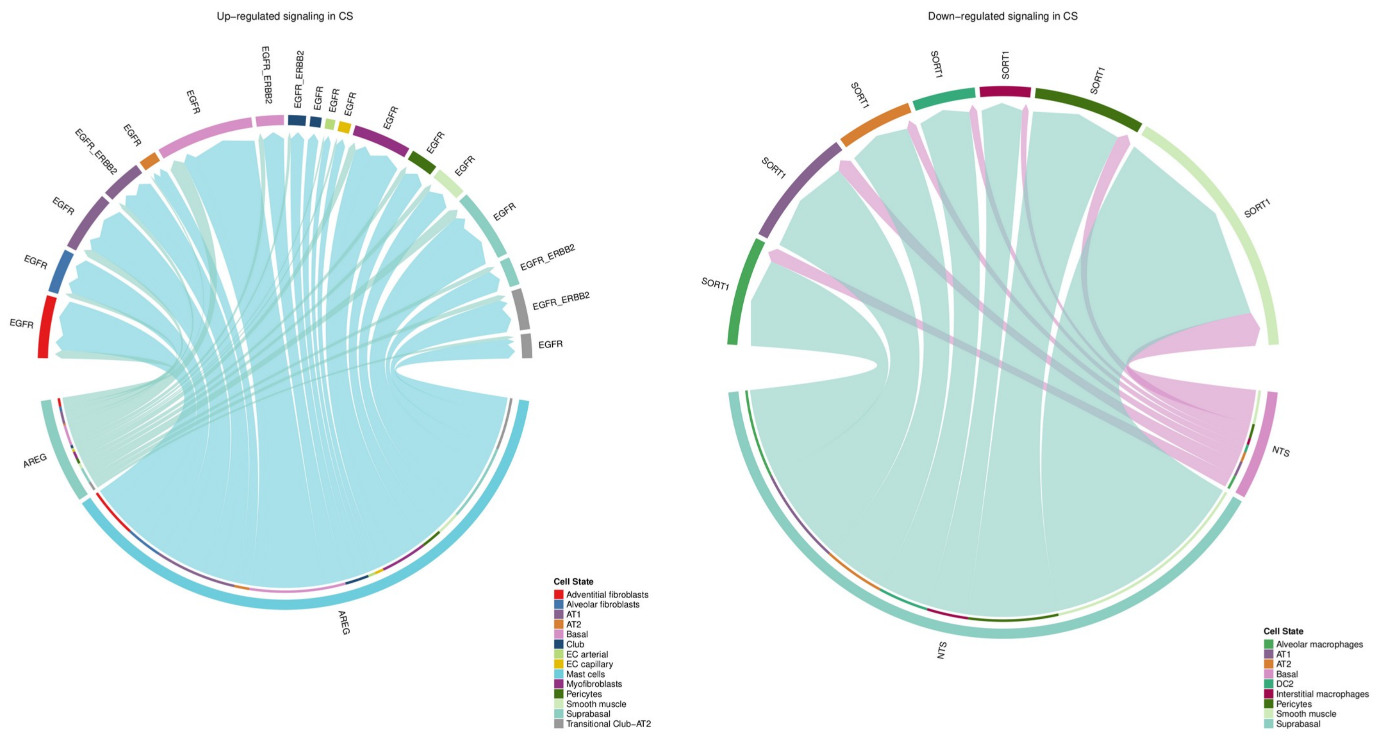


Figure S23. Chord diagrams for (A) the up-regulated signaling pathway EGF (Epidermal growth factor), and (B) the down-regulated signaling pathway NTS (Neurotensin).

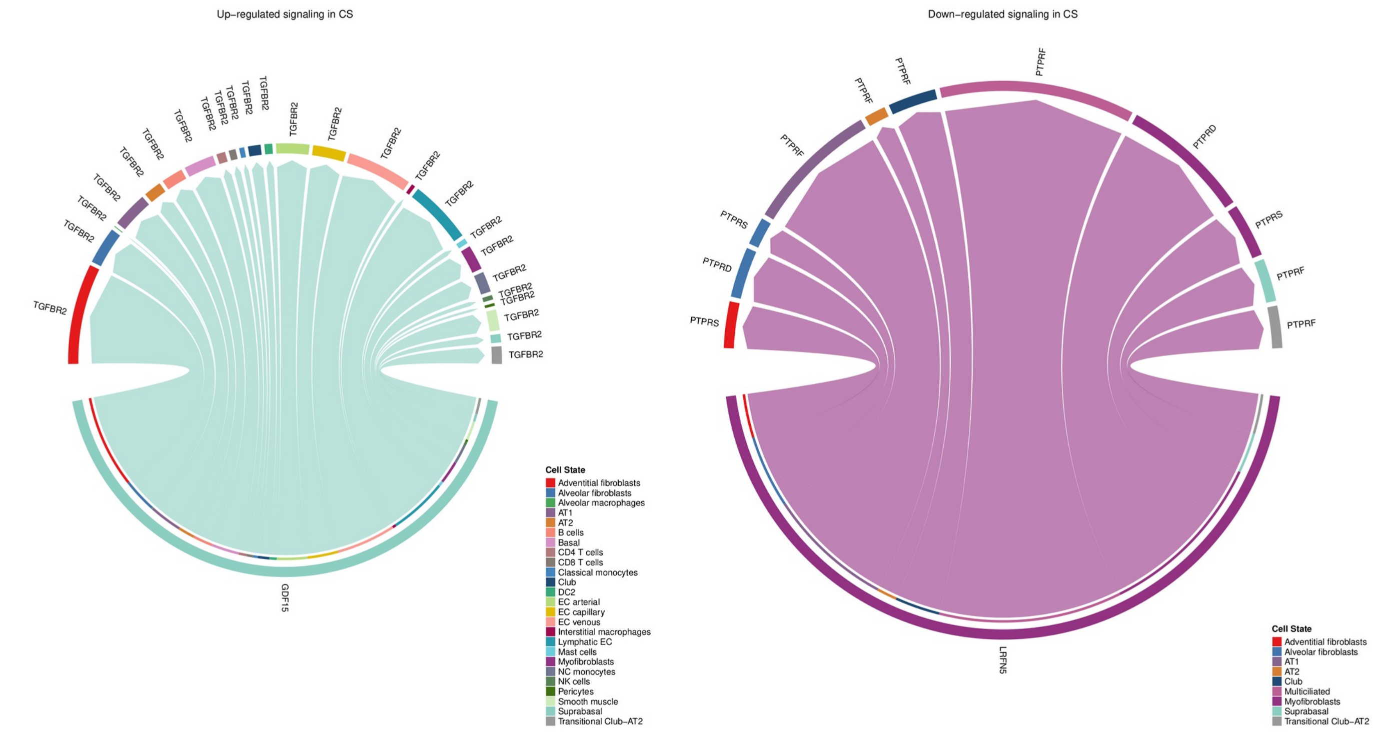


Figure S24. Chord diagrams for (A) the up-regulated signaling pathway GDF (Growth differentiation factor), and (B) the down-regulated signaling pathway PTPR (protein tyrosine phosphatase receptor).

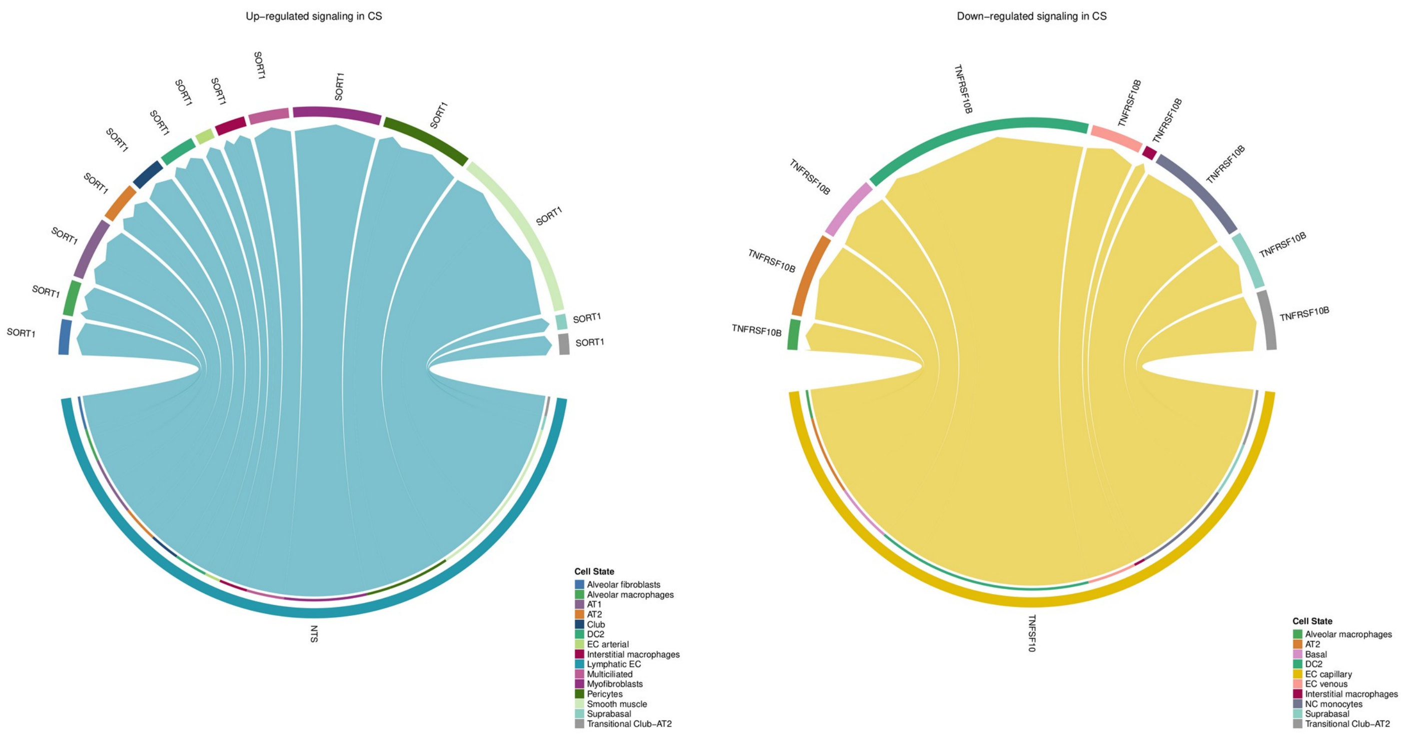


Figure S25. Chord diagrams for (A) the up-regulated signaling pathway NTS (Neurotensin), and (B) the down-regulated signaling pathway TRAIL (tumor necrosis factor-related apoptosis-inducing ligand).

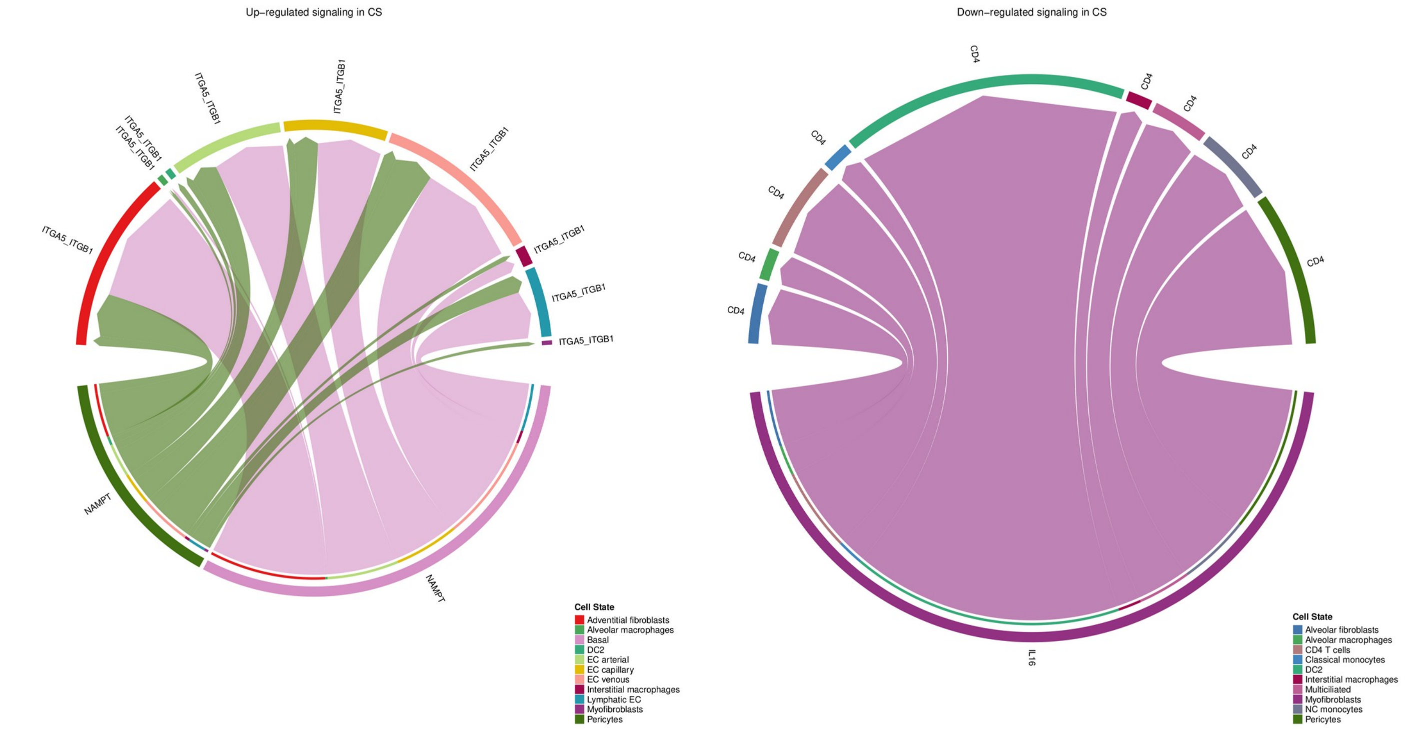


Figure S26. Chord diagrams for (A) the up-regulated signaling pathway VISFATIN, and (B) the down-regulated signaling pathway IL16.
